# Post-Receptor Dissociation of Estrogen Signaling in Macrophage-Infiltrated Meningiomas: A Multi-Method Deconvolution Study of 968 Transcriptomes

**DOI:** 10.64898/2026.05.19.26353588

**Authors:** Daniele Piccolo, Marco Vindigni

## Abstract

**Background:** Meningiomas exhibit well-established hormonal biology, yet no study has examined whether myeloid immune infiltration interacts with estrogen-responsive transcription in this tumor type.

**Methods:** We applied three-method consensus immune deconvolution (EPIC, MCPcounter, CIBERSORTx) to 968 harmonized meningioma RNA-seq transcriptomes from five public datasets, stratified by Thirimanne et al. (2024) transcriptomic subtypes. Competitive gene set enrichment compared macrophage-high versus macrophage-low tertiles with sex-adjusted, purity-adjusted, and method-independent sensitivity analyses. Survival modeling tested both total macrophage burden and a decomposed microglia-to-macrophage ratio validated against single-cell ground truth (pseudo-bulk r = 0.77).

**Results:** Macrophage-high tumors showed significant suppression of estrogen response gene sets (FDR = 4.9 × 10^−5^) despite paradoxical ESR1 upregulation (log_2_FC = +0.40, FDR = 2.5 × 10^−26^) and PGR downregulation (log_2_FC = −0.34, FDR = 2.7 × 10^−3^), indicating post-receptor transcriptional disruption. This signal strengthened after sex adjustment (FDR = 1.9 × 10^−6^) and was confirmed across a multi-layer sensitivity battery (eleven analyses including reference-matrix-independent, purity-adjusted, rotation-based self-contained, and empirical-null tests; all FDR < 3 × 10^−4^ in the relevant convergent tests).

Myeloid infiltration was strongly subtype-dependent (Kruskal–Wallis p = 7.4 × 10^−16^) but grade-independent (p = 0.399), with CSF1R enriched in the macrophage-dominant Cluster B. Neither total macrophage score (HR = 0.90, p = 0.53; N = 102) nor a decomposed microglia/macrophage ratio (HR = 0.92, p = 0.46; N = 101) predicted recurrence-free survival.

**Conclusions:** The pre-registered primary endpoint — macrophage infiltration score predicting recurrence-free survival — was not supported; the estrogen–immune dissociation emerged from secondary exploratory gene-set analysis and requires independent validation. Macrophage-infiltrated meningiomas exhibit a previously unreported dissociation between maintained ESR1 expression and suppressed estrogen-responsive transcription, with implications for hormonal therapy stratification.

## INTRODUCTION

Meningiomas are the most prevalent primary intracranial tumors, accounting for approximately 40% of all central nervous system neoplasms diagnosed annually [21]. Their hormonal biology is among the most established of any CNS tumor: progesterone receptors (PR) are expressed in approximately 70% of WHO grade I tumors, declining with increasing grade and serving as a favorable prognostic marker [6, 23]. The female-to-male incidence ratio is approximately 2:1, and exogenous progestin exposure, including cyproterone acetate, chlormadinone acetate, and nomegestrol acetate, is associated with meningioma growth [4, 21]. This hormonal dependence has motivated therapeutic exploration of anti-progestational agents, most notably mifepristone, though clinical trials have yielded inconsistent results, and no hormonal therapy has gained regulatory approval for meningioma [8]. Estrogen receptor expression in meningioma is more complex: while ERα (ESR1) is immunohistochemically detectable in a subset of tumors, it is not the dominant hormonal driver, and the clinical significance of estrogen signaling in meningioma remains incompletely understood [3, 23]. The 2021 WHO classification substantially revised meningioma grading by incorporating molecular parameters — CDKN2A/B deletion, TERT promoter mutation, and BAP1 loss — reflecting the heterogeneous biological behavior poorly captured by histological grade alone [17].

Independent of hormonal biology, the tumor microenvironment (TME) has emerged as a critical determinant of meningioma biology. Tumor-associated macrophages (TAMs) constitute the dominant immune population, comprising approximately 18–40% of all cells with predominantly M2 (immunosuppressive) polarization [9, 24]. Recent large-scale studies have substantially advanced understanding of meningioma myeloid biology. Maas et al. (2026) defined a microenvironment-determined risk continuum across 4,502 methylomes, demonstrating that NF2-mutant meningioma outcomes are driven by the shift from brain-resident microglia to peripheral myeloid-derived macrophages rather than total myeloid burden, validated by immunohistochemistry (PU.1 staining; HR = 2.00, 95% CI 1.45–2.76, N = 1,378) [16]. Lötsch et al. (2025) independently demonstrated that M2-like TAM density is an independent predictor of poor progression-free survival (HR = 2.11, P = 0.023, N = 680) [15]. Guo et al. (2026) profiled the immune microenvironment of 2,727 meningiomas using multi-modal approaches, reporting that macrophages constitute approximately 40% of tumor mass (IQR 27–60%) with the shift being qualitative (toward M2 polarization) rather than quantitative across grades [9]. Fan et al. (2025) revealed macrophage diversity, rather than total burden, as a key determinant of prognosis using integrated multi-omics clustering of 302 paired meningioma samples [7]. Thirimanne et al. (2024) assembled the largest meningioma RNA-seq meta-cohort to date (N = 1,298 samples from 13 datasets), defining seven transcriptomic subtypes (clusters A–G) with distinct molecular features and recurrence rates [29], but did not stratify immune infiltration by these clusters, leaving the question of subtype-specific immune architecture unanswered.

These studies collectively established myeloid composition as prognostically relevant in meningioma, yet none examined its intersection with the tumor’s hormonal transcriptional landscape. This gap is notable because immune–endocrine crosstalk is increasingly recognized in other malignancies: TAMs suppress ERβ expression via PI3K/AKT/FOXO3a in triple-negative breast cancer [32], while macrophage-derived IL-17A upregulates ERα through epigenetic mechanisms in endometrial cancer [20]. Whether similar interactions occur in meningioma, a hormone-responsive tumor with a macrophage-dominant microenvironment, has not been investigated.

We designed a systematic multi-method immune deconvolution study of 968 meningioma transcriptomes to address this gap. Our objectives were: (1) to determine whether myeloid infiltration patterns are organized by transcriptomic subtype or WHO grade; (2) to characterize the relationship between macrophage infiltration and estrogen-responsive transcriptional programs; and (3) to evaluate the CSF1R/TAM axis across transcriptomic subtypes as a candidate for subtype-stratified immunotherapy.

## METHODS

### Data Sources and Batch Harmonization

We assembled a meningioma transcriptomic meta-cohort from five publicly available GEO datasets: GSE212666 (HKU/UCSF; N = 302), GSE252291 (UW/FHCC; N = 279), GSE183653 (UCSF 2022; N = 185), GSE136661 (Baylor; N = 160), and GSE101638 (UCSF 2018; N = 42), totaling 968 samples across four institutions (Table 1). Expression data were provided in mixed formats (log_2_-transformed TPM, raw TPM, FPKM, or HTSeq counts), all converted to log_2_(normalized expression + 1) units prior to integration. Two initially identified accessions were excluded: GSE183656 (DNA methylation IDAT arrays) and GSE139652 (ChIP-seq BigWig files). WHO grade distribution in the Oncoscape clinical metadata (N = 999) was: grade I 58.8% (N = 588), grade II 31.0% (N = 310), grade III 6.5% (N = 65), and unknown 3.7% (N = 37).

**Table 1.**
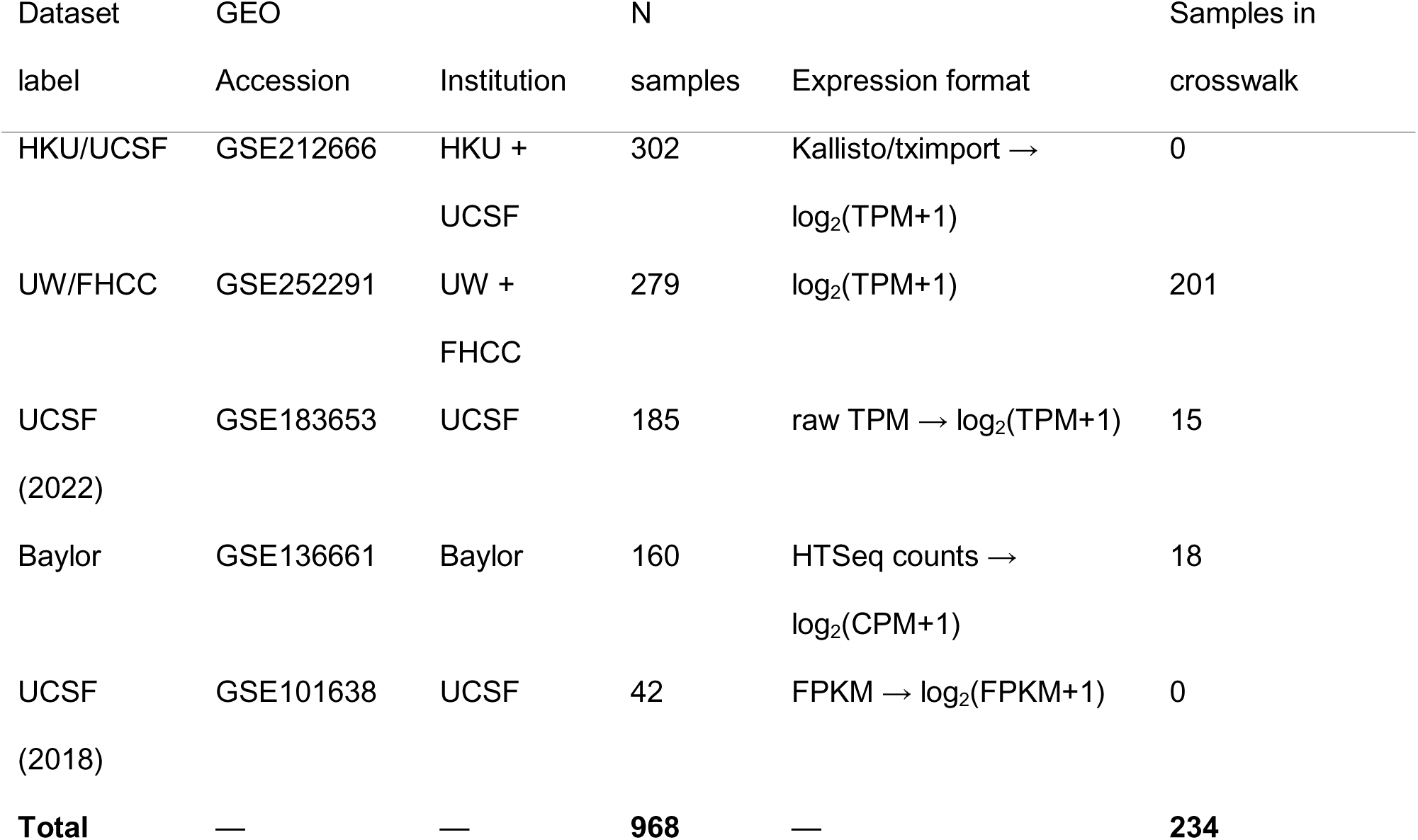
Dataset composition of the harmonized expression matrix (N = 968 samples).

| Dataset | GEO |  | N |  | Samples in |
| --- | --- | --- | --- | --- | --- |
| label | Accession | Institution | samples | Expression format | crosswalk |
| HKU/UCSF | GSE212666 | HKU + | 302 | Kallisto/tximport → | 0 |
|  |  | UCSF |  | log<sub>2</sub>(TPM+1) |  |
| UW/FHCC | GSE252291 | UW + | 279 | log<sub>2</sub>(TPM+1) | 201 |
|  |  | FHCC |  |  |  |
| UCSF | GSE183653 | UCSF | 185 | raw TPM → log<sub>2</sub>(TPM+1) | 15 |
| (2022) |  |  |  |  |  |
| Baylor | GSE136661 | Baylor | 160 | HTSeq counts → | 18 |
|  |  |  |  | log<sub>2</sub>(CPM+1) |  |
| UCSF | GSE101638 | UCSF | 42 | FPKM → log<sub>2</sub>(FPKM+1) | 0 |
| (2018) |  |  |  |  |  |
| **Total** | — | — | **968** | — | **234** |

ComBat-seq was not applicable because expression data were provided in mixed formats (TPM, FPKM, and counts); limma::removeBatchEffect [26], which operates on continuous normalized values, was therefore used on the log_2_-normalized expression matrix. The harmonized matrix contained 17,585 genes × 968 samples; 1,277,357 values (7.5% of total) were clipped from negative to zero, consistent with expected behavior of the limma shift-and-mean method. Batch correction adequacy was assessed by PERMANOVA (vegan::adonis2) on the top 500 most variable genes: batch-explained variance decreased from R^2^ = 0.663 to R^2^ = 0.0004 (pre-specified threshold R^2^ < 0.05), while WHO grade signal was preserved (R^2^ = 0.068 → 0.062, p = 0.002 before and after). Principal component analysis was performed before and after correction, colored by batch identity and WHO grade independently (Supplementary Fig. S1).

### Immune Deconvolution

Three computational immune deconvolution methods were applied to the batch-corrected expression matrix. EPIC (Estimating the Proportions of Immune and Cancer cells) [25] was run using the TRef reference matrix in absolute mode via the R package omnideconv. Fourteen samples with non-converging optimization were retained with a cautionary note. MCPcounter (Microenvironment Cell Populations counter) [1] was applied via omnideconv to quantify eight leukocyte and stromal populations. CIBERSORTx [19] was run in absolute mode using the LM22 22-cell reference matrix through the licensed web portal (https://cibersortx.stanford.edu); 279/968 samples (28.8%) achieved statistical significance at p < 0.05. CIBERSORTx macrophage fractions were defined as the sum of LM22 M0, M1, and M2 monocyte/macrophage subtypes.

For each homologous cell-type pair across methods, pairwise Spearman correlation was computed on all N = 968 samples. A cell type was classified as concordant if at least 2 of 3 method pairs achieved r ≥ 0.50 (pre-specified protocol threshold). A composite macrophage infiltration score (macro_score) was computed as the mean of within-method z-scored EPIC macrophage, MCPcounter monocytic lineage, and CIBERSORTx macrophage fractions. The primary endpoint cell type (EPIC macrophages) exhibited a fully continuous distribution with no samples at the zero floor (range: 0.000–0.248; Q1 = 0.025, Q3 = 0.064), confirming that batch correction clipping did not extinguish low-abundance immune signal.

CIBERSORTx estimates for samples not reaching significance (71.2%) were retained in the composite score to avoid selection bias; the 2-method composite (EPIC + MCPcounter) yielded identical cluster rankings (r = 1.00), confirming that CIBERSORTx did not drive primary results. For cell types estimated by only 2 of 3 methods (CAFs, endothelial cells), the concordance rule was adapted to require 1/1 available pair above threshold. Per-cell-type concordance values and CIBERSORTx p < 0.05 subset analyses are in Supplementary Table S2.

### Transcriptomic Subtype Linkage

Thirimanne et al. (2024) transcriptomic cluster assignments (A–G) [30] were linked to the expression matrix by direct sample-ID matching (UW/FHCC, N = 201), probabilistic matching using age/sex/grade (Baylor, N = 18), and M-number matching (UCSF 2022, N = 15), yielding N = 234 cluster-linked samples. Crosswalk methodology details are in Supplementary Methods.

### Differential Expression and Gene Set Enrichment

Differential expression between macrophage tertile-high (N = 323) and tertile-low (N = 323) samples was performed using limma [26] on the log_2_-normalized batch-corrected matrix, with batch as covariate. Tertile boundaries were defined by CIBERSORTx macrophage scores (Q33 = 0.5064, Q66 = 0.7442). Genes with mean log-CPM > 0.5 were retained (13,240 genes tested). Significance thresholds: FDR < 0.05, |log_2_FC| > 0.5. To assess purity confounding, EPIC otherCells fraction (range 0–0.949; median 0.563) was added as a continuous covariate in a sensitivity analysis.

Gene set enrichment was performed using camera() [31] from the limma package applied to 50 MSigDB Hallmark gene sets [14] (minimum 10 members per set), with inter-gene correlation fixed at 0.01 (recommended for RNA-seq data). FDR was computed using the Benjamini–Hochberg procedure across the 50 Hallmark sets within each camera analysis independently. Four independent analyses were performed: (1) primary (CIBERSORTx tertile, N = 646); (2) sex-adjusted (sex as design matrix covariate, N = 79 with documented biological sex — 43 female, 36 male, predominantly from UW/FHCC); (3) purity-adjusted (EPIC otherCells fraction as covariate, N = 646); and (4) MCPcounter-based (MCPcounter monocytic lineage tertile as the independent stratification variable, precluding overlap with CIBERSORTx/LM22 reference). The MCPcounter-based analysis also served as the primary differential expression result for CSF1R assessment, with the CIBERSORTx-based analysis as confirmatory, to avoid circularity with the LM22 reference matrix. As these analyses test the same hypothesis on overlapping samples using different analytical specifications, they represent sensitivity analyses rather than independent tests; formal correction across analyses was not applied. The primary analysis (CIBERSORTx tertile, FDR = 4.9 × 10^−5^) was pre-specified. The full robustness battery comprises eleven analyses: (1) primary CIBERSORTx tertile; (2) continuous macrophage score (N = 968); (3) within Cluster A; (4) within Cluster B; (5) formal cluster × macrophage interaction term; (6) leave-one-dataset-out across five datasets (collapsed as a single sensitivity family); (7) sex adjustment; (8) EPIC-otherCells purity adjustment; (9) meningothelial-lineage purity proxy (PTGDS/CLDN1/SSTR2 composite); (10) self-contained rotation-based fry test (independent of inter-gene correlation assumption); (11) empirical null from 1,000 random size-matched gene sets. An MCPcounter-based reference-matrix-independent confirmation was additionally performed. Partial-correlation analyses of ESR1/PGR/ESR2 with the continuous macrophage score were conducted in the N = 968 cohort with batch and purity (EPIC otherCells or meningothelial proxy) as adjustment covariates.

### Therapeutic Target Profiling

Eight pre-specified therapeutic targets (CSF1R, CSF1, IL34, CD274, CDK4, CDK6, VEGFA, PDGFRA) were profiled by expression level across Thirimanne clusters using Kruskal–Wallis tests with Benjamini–Hochberg FDR correction (N = 225). ssGSEA pathway scores (CSF1R_TAM, Immune_Activation, Cell_Cycle, PI3K_AKT) were computed using GSVA [10] in single-sample mode.

### Microglia/Macrophage Ratio and Single-Cell Validation

To test whether decomposing the myeloid compartment into microglia-like and macrophage-like subpopulations rescues prognostic signal, we constructed expanded gene sets anchored to the Maas et al. core markers [16]: a microglia set (15 genes: TMEM119, P2RY12, P2RY13, GPR34, SLC2A5, CX3CR1, HEXB, SALL1, CSF1R, TREM2, OLFML3, SELPLG, SPI1, SIGLEC8, AIF1) and a macrophage set (17 genes: C3, F10, EMILIN2, F5, GDA, HP, SPP1, CD163, MARCO, FOLR2, MSR1, CD68, ITGAM, CCL2, TGFB1, IL10, CD83). Per-sample ssGSEA scores were computed using GSVA [10], and the microglia-to-macrophage ratio was defined as ssGSEA_microglia_ − ssGSEA_macrophage_ (z-scored). The ratio was validated against single-cell ground truth using pseudo-bulk profiles from the Maas snRNA-seq cohort (44,266 nuclei, 25 evaluable samples, 15,779 TAMs) [16]. Full gene set construction rationale, circularity analysis, and sensitivity analyses are described in Piccolo and Vindigni (2026) [22].

### Survival Analysis

Recurrence-free survival (RFS) was analyzed in two complementary models. For total macrophage burden, a restricted LASSO-Cox model was specified with five pre-selected terms (macro_score, T-cell score, CSF1R pathway score, WHO grade, macrophage × subtype interaction) in N = 102 patients with 73 events (median follow-up 71.0 months); LASSO penalization retained only WHO grade, yielding a 2-term multivariable model (EPV = 36.5). For the decomposed microglia/macrophage ratio, univariable and multivariable Cox regression (adjusted for WHO grade) were performed in N = 101 patients with 73 events (median follow-up 110.2 months) [22]. Permutation testing (10,000 permutations) assessed the ratio’s empirical significance. Proportional hazards assumptions were verified by Schoenfeld residual tests [27]. Statistical power was assessed via the Schoenfeld approximation (two-sided α = 0.05, 80% power); under these assumptions, 73 events correspond to a minimum detectable hazard ratio of 1.40 (0.71) per 1-SD increase in a continuous covariate and 1.93 (0.52) for a balanced binary contrast. An exploratory NF2 expression proxy analysis stratified the survival cohort by median NF2 mRNA expression [22]. As a pre-specified sensitivity analysis, a Fine–Gray subdistribution hazards model was fitted for the primary macrophage score endpoint to address potential competing-risks bias; because no competing deaths were separable from the pooled recurrence endpoint in the available metadata, the Fine–Gray and cause-specific Cox models coincide.

All analyses were conducted in R version 4.5.3 All stochastic analyses used set.seed(42).

## RESULTS

### Cohort and Batch Correction

The harmonized expression matrix comprised 968 meningioma RNA-seq samples from five GEO datasets spanning four institutions (Table 1). WHO grade distribution in the Oncoscape clinical metadata (N = 999) was: grade I 58.8% (N = 588), grade II 31.0% (N = 310), grade III 6.5% (N = 65), and unknown 3.7% (N = 37). Batch correction with limma::removeBatchEffect reduced dataset-explained variance from 66.3% to 0.04% (PERMANOVA R^2^: 0.663 → 0.0004) while preserving WHO grade signal (R^2^ = 0.068 → 0.062, p = 0.002 before and after; Supplementary Fig. S1).

### Myeloid Infiltration Is Organized by Transcriptomic Subtype, Not WHO Grade

Of seven cell types assessed by three-method consensus, only macrophages/monocytes (EPIC × MCPcounter r = 0.764; EPIC × CIBERSORTx r = 0.618; MCPcounter × CIBERSORTx r = 0.773), cancer-associated fibroblasts (r = 0.818), and endothelial cells (r = 0.703) reached cross-method concordance; all four lymphoid populations were non-concordant (r ≤ 0.11; Supplementary Table S2).

In the 234 samples with Thirimanne transcriptomic cluster assignments (cluster A = 82, B = 39, C = 46, D = 37, E = 11, F = 8, G = 2), macrophage infiltration scores differed markedly across subtypes (Kruskal–Wallis χ^2^= 77.02, df = 4, p = 7.4 × 10^−16^; clusters A–E; F [N = 8] and G [N = 2] excluded from test due to insufficient sample size) but not across WHO grades (χ^2^= 1.84, df = 2, p = 0.399; Fig. 1A–B). Post-hoc Dunn analysis identified Cluster B as the macrophage-enriched subtype and Cluster C as the immune desert (B vs C: Z = 7.93, q = 2.1 × 10^−14^; B vs A: q = 1.3 × 10^−5^; B vs D: q = 1.1 × 10^−8^; B vs E: q = 2.8 × 10^−7^; Fig. 1C; Supplementary Table S3). CAF and endothelial scores replicated this pattern (cluster p = 2.7 × 10^−14^ and p = 5.1 × 10^−6^, respectively; grade p > 0.25 for both), demonstrating that the TME phenotype is shaped by transcriptomic identity rather than histological grade.

**Fig. 1.**
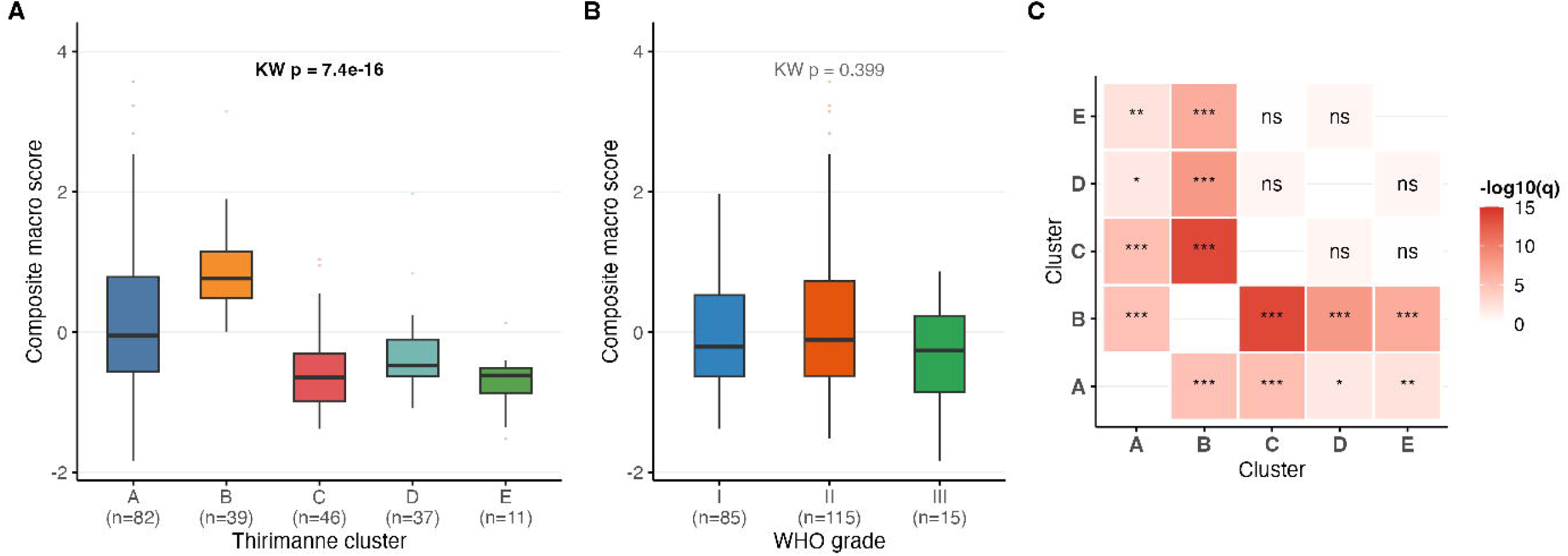
Macrophage infiltration is organized by transcriptomic subtype, not WHO grade. (A) Boxplot of macro_score by Thirimanne cluster (A–E; N = 234 linked samples). Kruskal–Wallis p = 7.4 × 10^−16^. Cluster B = macrophage-enriched; Cluster C = immune desert. Significant Dunn pairwise comparisons indicated (BH-FDR q values). (B) Boxplot of macro_score by WHO grade (I–III; same N = 234). Kruskal–Wallis p = 0.399 (not significant). (C) Dunn post-hoc matrix for macrophage scores across clusters; color indicates −log_10_(q).

Critically, in the same 234 cluster-linked samples, all of which have WHO grade annotations (Grade I = 92, Grade II = 125, Grade III = 17), macrophage score did not differ across grades (mean: Grade I = −0.04, Grade II = +0.04, Grade III = −0.27; p = 0.399), confirming that the cluster effect and grade-independence are demonstrated in the same population, not inferred from disjoint patient sets. This was independently confirmed in the full Baylor cohort (N = 160; KW χ^2^ = 2.544, df = 2, p = 0.280). As a pre-specified sensitivity analysis, restricting the Kruskal–Wallis test to the UW/FHCC cohort alone (N = 193, the subset with direct sample-ID matching) yielded H = 75.21, p = 3.47 × 10^−14^, with identical cluster rank ordering, confirming that subtype-dependent immune stratification is not attributable to the smaller probabilistically matched contributions.

### Macrophage-Infiltrated Tumors Show Suppression of Estrogen-Responsive Transcription Despite Maintained ESR1 Expression

Competitive gene set enrichment (camera) comparing macrophage-high (N = 323) versus macrophage-low (N = 323) tumors identified 11 significantly enriched Hallmark gene sets (FDR < 0.05). Upregulated sets reflected a pro-inflammatory myeloid signature: ALLOGRAFT_REJECTION (FDR = 1.76 × 10^−12^), INTERFERON_GAMMA_RESPONSE (FDR = 8.76 × 10^−8^), COMPLEMENT (FDR = 2.43 × 10^−6^), IL6_JAK_STAT3_SIGNALING (FDR = 5.24 × 10^−6^), and INFLAMMATORY_RESPONSE (FDR = 1.46 × 10^−5^). Unexpectedly, both ESTROGEN_RESPONSE_LATE (FDR = 2.79 × 10^−5^) and ESTROGEN_RESPONSE_EARLY (FDR = 4.89 × 10^−5^) were significantly downregulated in macrophage-high tumors (Fig. 2A–B).

**Fig. 2.**
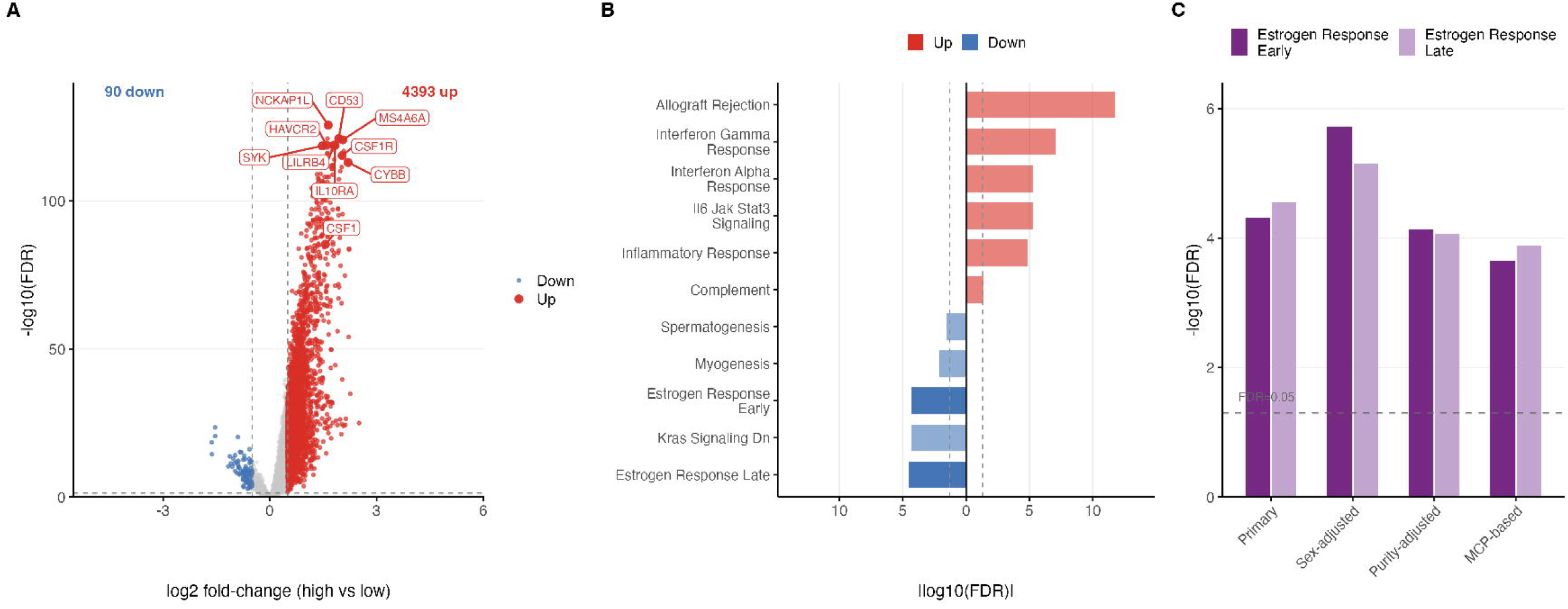
Macrophage-infiltrated tumors exhibit an inverse estrogen–immune axis. (A) Volcano plot of limma differential expression (N = 13,240 genes; CIBERSORTx macrophage tertile high vs low, N = 646). CSF1R and top myeloid genes annotated. (B) Camera barcode plot for Hallmark gene sets (FDR < 0.05). Immune activation sets (top) upregulated; estrogen response sets (bottom) downregulated in macrophage-high tumors. (C) Individual hormone receptor expression by macrophage tertile: ESR1 upregulated (log_2_FC = +0.40), PGR downregulated (log_2_FC = −0.34), ESR2 unchanged — demonstrating post-receptor disruption rather than receptor silencing. (D) Comparison of ESTROGEN_RESPONSE_EARLY FDR across the sensitivity battery (Table 2). Sex-adjusted FDR (1.91 × 10^−6^) is stronger than primary, confirming the signal is not sex-driven.

**Table 2.**
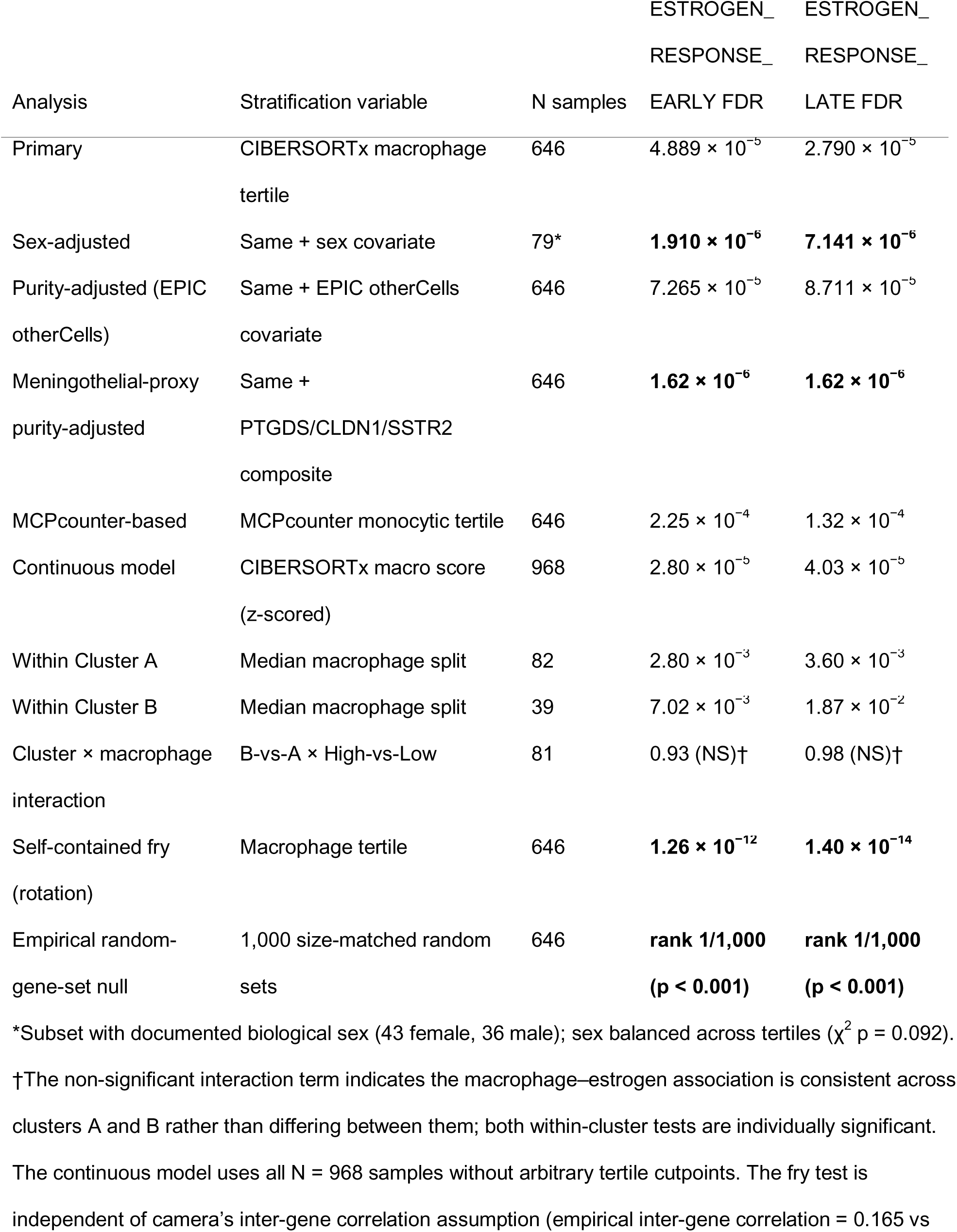
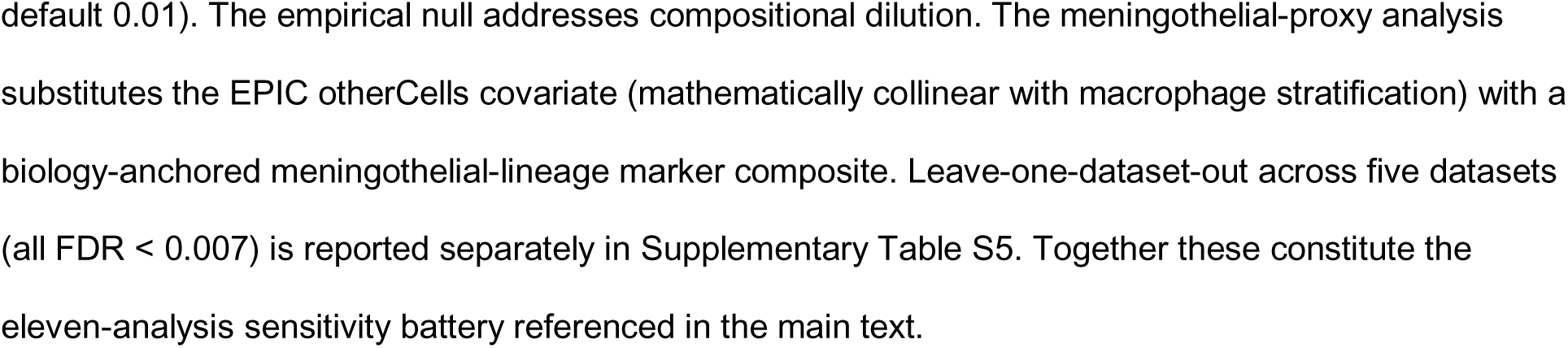
Estrogen response gene set FDR values across the eleven-analysis sensitivity battery.

| Analysis | Stratification variable | N samples | ESTROGEN_<br>RESPONSE_<br>EARLY FDR | ESTROGEN_<br>RESPONSE_<br>LATE FDR |
| --- | --- | --- | --- | --- |
| Primary | CIBERSORTx macrophage<br>tertile | 646 | $4.889 \times 10^{-5}$ | $2.790 \times 10^{-5}$ |
| Sex-adjusted | Same + sex covariate | 79* | **$1.910 \times 10^{-6}$** | **$7.141 \times 10^{-6}$** |
| Purity-adjusted (EPIC<br>otherCells) | Same + EPIC otherCells<br>covariate | 646 | $7.265 \times 10^{-5}$ | $8.711 \times 10^{-5}$ |
| Meningothelial-proxy<br>purity-adjusted | Same +<br>PTGDS/CLDN1/SSTR2<br>composite | 646 | **$1.62 \times 10^{-6}$** | **$1.62 \times 10^{-6}$** |
| MCPcounter-based | MCPcounter monocytic tertile | 646 | $2.25 \times 10^{-4}$ | $1.32 \times 10^{-4}$ |
| Continuous model | CIBERSORTx macro score<br>(z-scored) | 968 | $2.80 \times 10^{-5}$ | $4.03 \times 10^{-5}$ |
| Within Cluster A | Median macrophage split | 82 | $2.80 \times 10^{-3}$ | $3.60 \times 10^{-3}$ |
| Within Cluster B | Median macrophage split | 39 | $7.02 \times 10^{-3}$ | $1.87 \times 10^{-2}$ |
| Cluster × macrophage<br>interaction | B-vs-A × High-vs-Low | 81 | 0.93 (NS)† | 0.98 (NS)† |
| Self-contained fry<br>(rotation) | Macrophage tertile | 646 | **$1.26 \times 10^{-12}$** | **$1.40 \times 10^{-14}$** |
| Empirical random-gene-set null | 1,000 size-matched random<br>sets | 646 | **rank 1/1,000**<br>**(p < 0.001)** | **rank 1/1,000**<br>**(p < 0.001)** |

\*Subset with documented biological sex (43 female, 36 male); sex balanced across tertiles ( $\chi^2$ p = 0.092).
†The non-significant interaction term indicates the macrophage–estrogen association is consistent across
clusters A and B rather than differing between them; both within-cluster tests are individually significant.
The continuous model uses all N = 968 samples without arbitrary tertile cutpoints. The fry test is
independent of camera's inter-gene correlation assumption (empirical inter-gene correlation = 0.165 vs

Examination of individual hormone receptor genes revealed a paradoxical dissociation. ESR1 was significantly upregulated in macrophage-high tumors (log_2_FC = +0.40, FDR = 2.48 × 10^−26^), while PGR, the dominant hormone receptor in meningioma, was significantly downregulated (log_2_FC = −0.34, FDR = 2.70 × 10^−3^). ESR2 was unchanged (log_2_FC = −0.06, FDR = 0.19). This pattern was confirmed in the independent MCPcounter-based analysis (ESR1 log_2_FC = +0.44, FDR = 1.34 × 10^−25^; PGR log_2_FC = −0.42, FDR = 1.65 × 10^−4^). The estrogen response suppression therefore reflects disruption of downstream transcriptional programs in macrophage-infiltrated tumors despite maintained receptor expression, pointing to post-receptor signaling interference rather than receptor silencing (Fig. 2C). Consistent with partial macrophage contribution to bulk ESR1 signal, the marginal Spearman correlation of ESR1 with the macrophage consensus score (ρ = 0.54) attenuated after adjustment for purity and batch (partial β = +0.136, p = 0.014 with EPIC otherCells; β = +0.176, p = 4.6 × 10^−4^ with the meningothelial-lineage proxy). PGR expression remained strongly inversely associated with macrophage infiltration after identical adjustment (partial β = −0.73, p = 0.008; β = −0.89, p = 4.0 × 10^−4^), indicating that the PGR downregulation is not explained by purity or composition.

The signal was robust across a multi-layer sensitivity battery (eleven analyses including reference-matrix-independent, purity-adjusted, rotation-based self-contained, and empirical-null tests; Table 2). Sex adjustment in 79 sex-annotated samples (43 female, 36 male; χ^2^ p = 0.092 for sex balance across tertiles) strengthened rather than attenuated the signal (ESTROGEN_RESPONSE_EARLY FDR = 1.91 × 10^−6^; LATE FDR = 7.14 × 10^−6^), inconsistent with sex as the sole driver of the observed association, though the small subsample size limits definitive conclusions about signal amplification. Purity adjustment (EPIC otherCells fraction covariate) and the fully independent MCPcounter-based analysis confirmed significance (all FDR < 3 × 10^−4^; Table 2).

To address potential confounding by transcriptomic subtype and NF2 status, we performed within-cluster camera analyses. Within Cluster A (N = 82, median macrophage split: 41 high vs 41 low), both estrogen response gene sets remained significantly downregulated in macrophage-high tumors (EARLY FDR = 2.8 × 10^−3^; LATE FDR = 3.6 × 10^−3^). Within Cluster B (N = 39), the signal was likewise significant (EARLY FDR = 7.0 × 10^−3^; LATE FDR = 1.9 × 10^−2^). A formal cluster × macrophage interaction term was non-significant for both gene sets (EARLY interaction FDR = 0.93; LATE FDR = 0.98), indicating that the estrogen–immune dissociation operates consistently across both the NF2-mixed hormonally-competent Cluster A and the NF2-loss macrophage-enriched Cluster B, rather than being confined to a single transcriptomic subtype.

Three additional robustness tests confirmed the signal. A self-contained rotation-based gene set test (fry) yielded EARLY FDR = 1.26 × 10^−12^ and LATE FDR = 1.40 × 10^−14^, eliminating the possibility that the camera result depended on the fixed inter-gene correlation default (empirical inter-gene correlation = 0.165). An empirical null constructed from 1,000 random 200-gene sets drawn from the tested gene universe ranked the observed estrogen set enrichment statistic at rank 1/1,000 for both EARLY and LATE sets (empirical p < 0.001), excluding compositional dilution as the generator of the signal. Finally, substituting the EPIC otherCells purity covariate, mathematically collinear with the macrophage stratification, with an independent meningothelial-lineage purity proxy (mean z-score of PTGDS, CLDN1, and SSTR2) preserved the signal (EARLY FDR = 1.62 × 10^−6^; LATE FDR = 1.62 × 10^−6^), confirming the association is not an artifact of purity-macrophage collinearity.

A continuous macrophage score model across all 968 samples, without tertile cutpoints, confirmed the association (EARLY FDR = 2.8 × 10^−5^; LATE FDR = 4.0 × 10^−5^). Leave-one-dataset-out analysis showed the signal survived all five dataset drops (all FDR < 0.007), with the weakest result upon removal of UW/FHCC (EARLY FDR = 6.7 × 10^−3^; Supplementary Table S5). Leading-edge analysis identified classical hormone-responsive genes (PGR, KRT19, MUC1, MLPH, KLK10) as drivers, supporting specificity of the estrogen response signal rather than generic compositional dilution.

### CSF1R/TAM Axis Is Enriched in the Macrophage-Dominant Subtype

In the CIBERSORTx-based differential expression (N = 323 vs 323), 4,393 upregulated and 90 downregulated genes were identified (FDR < 0.05, |log_2_FC| > 0.5). The 49:1 asymmetry toward upregulation is biologically expected, as macrophage infiltration contributes an additive transcriptome without proportional silencing of tumor-cell genes. Top upregulated genes were classical myeloid effectors: NCKAP1L (log_2_FC = 1.64, FDR = 2.4 × 10^−126^), ARHGAP30 (1.62, FDR = 9.1 × 10^−122^), CD53 (1.93, FDR = 9.1 × 10^−122^), MS4A6A (2.05, FDR = 2.3 × 10^−121^), and LILRB4 (1.80, FDR = 1.5 × 10^−119^).

The MCPcounter-based analysis (reference-matrix-independent) identified CSF1R as the 29th-ranked gene of 13,217 tested (log_2_FC = 2.23, FDR = 2.1 × 10^−145^). In the confirmatory CIBERSORTx-based analysis, CSF1R ranked 13th (log_2_FC = 2.03, FDR = 4.7 × 10^−116^). The convergence of both methods confirms that CSF1R association with macrophage infiltration is not a reference-matrix artifact. The ligand CSF1 and the alternative ligand IL34 similarly ranked among the top-associated genes.

Therapeutic target profiling across Thirimanne clusters confirmed concordant enrichment: Cluster B exhibited the highest median CSF1R transcript abundance (median 6.60 log-CPM vs Cluster A = 5.11, C = 4.71, D = 4.82, E = 3.84; Kruskal–Wallis p = 1.4 × 10^−14^) and the highest CSF1 ligand expression (median 6.31 vs Cluster C = 4.53; p = 9.4 × 10^−17^), consistent with active paracrine CSF1–CSF1R signaling in this macrophage-enriched subtype (Supplementary Table S1). ssGSEA pathway scores further confirmed that CSF1R_TAM pathway enrichment is strongly subtype-dependent (KW χ^2^ = 76.73, q = 3.4 × 10^−15^), with Cell_Cycle (q = 4.2 × 10^−8^), Immune_Activation (q = 4.4 × 10^−9^), and PI3K_AKT (q = 1.1 × 10^−5^) pathways also varying significantly.

### Total and Decomposed Myeloid Scores Do Not Predict Recurrence-Free Survival

In the exploratory survival cohort (N = 102, 73 events), total macrophage infiltration score was not associated with recurrence-free survival in univariable (HR = 0.904, 95% CI 0.660–1.237, p = 0.53) or multivariable analysis (HR = 1.004, 95% CI 0.725–1.390, p = 0.98; Table 3). WHO grade was the only significant predictor (univariable HR = 1.739, 95% CI 1.206–2.507, p = 0.003). In the LASSO-Cox model with five pre-specified terms, only WHO grade was retained at λ_min_ (Fig. 3A–B). Fine–Gray subdistribution hazards on the same patient set (N = 101, 73 events) yielded identical estimates (sHR = 1.00, 95% CI 0.78–1.27, p = 0.98).

**Fig. 3.**
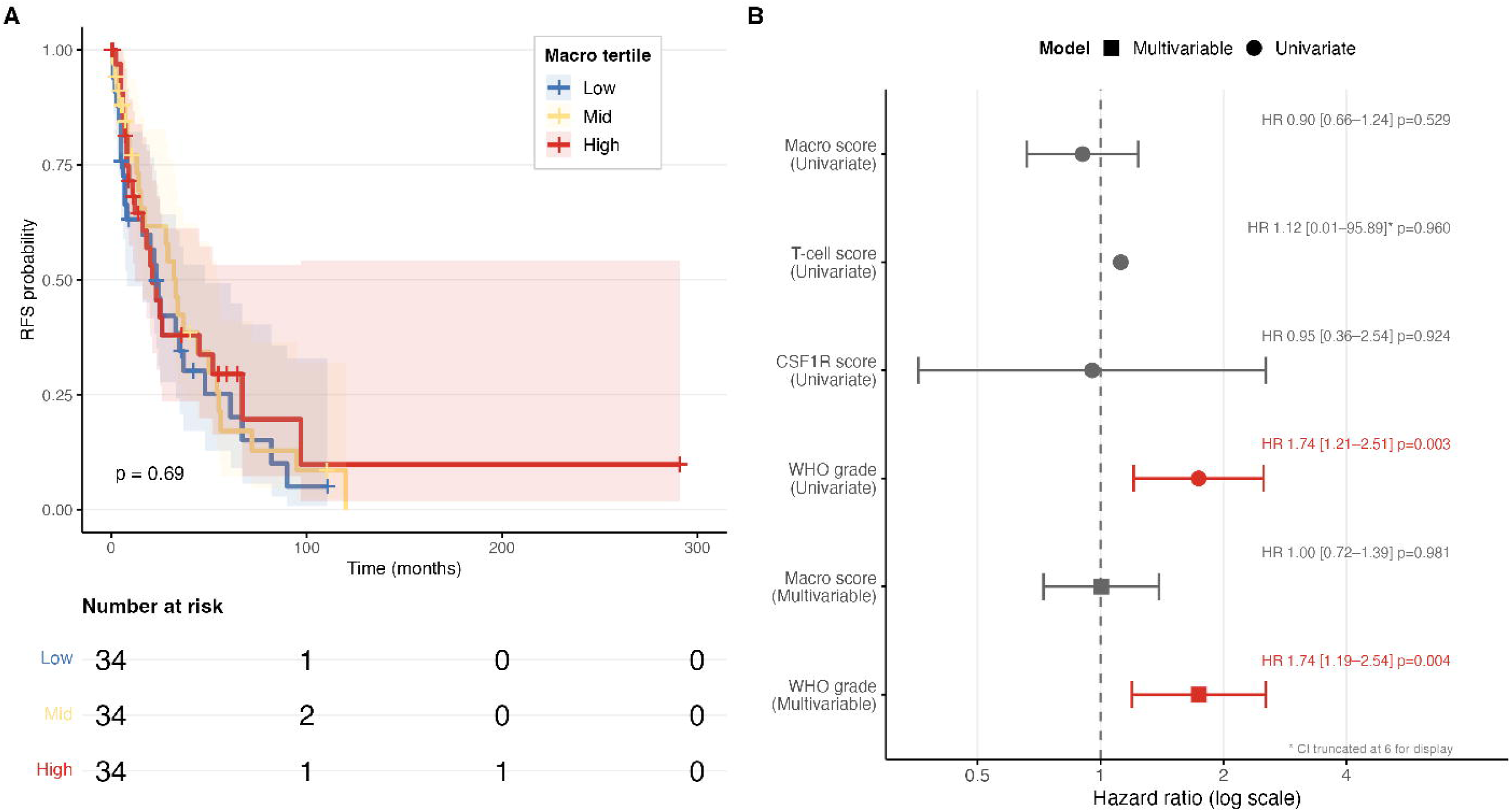
Total and decomposed myeloid scores do not predict recurrence-free survival. (A) Kaplan–Meier curves for RFS by macro_score tertile (high/mid/low; N = 102, 73 events). Three-group log-rank p = 0.69. (B) Forest plot of univariate Cox HR estimates for all tested predictors. WHO grade is the only significant predictor (HR = 1.74, p = 0.003); all immune scores non-significant. (C) Kaplan–Meier curves for RFS by microglia/macrophage ratio tertile (N = 101, 73 events). Log-rank p = 0.79 [22]. Neither total burden nor decomposed composition predicts RFS despite the ratio’s biological validity (pseudo-bulk r = 0.77 vs single-cell ground truth).

**Table 3.** Survival analysis results: total macrophage score and decomposed microglia/macrophage ratio.

| Analysis | Model | Term | HR | 95% CI | p-value | C-index |
| --- | --- | --- | --- | --- | --- | --- |
| **Total macrophage** |  |  |  |  |  |  |
| (N = 102, 73 events) | Univariable | macro_score | 0.904 | 0.660–1.237 | 0.53 | 0.576 |
|  | Univariable | WHO grade | **1.739** | **1.206–2.507** | **0.003** | — |
|  | Multivariable | macro_score | 1.004 | 0.725–1.390 | 0.98 | — |
|  | Multivariable | WHO grade | **1.741** | **1.193–2.540** | **0.004** | — |
| **Decomposed ratio** |  |  |  |  |  |  |
| (N = 101, 73 events) | Univariable | Ratio (per SD) | 0.92 | 0.72–1.16 | 0.46 | 0.551 |
|  | Univariable | WHO grade | **1.74** | **1.21–2.51** | **0.003** | 0.594 |
|  | Multivariable | Ratio (per SD) | 0.98 | 0.77–1.24 | 0.85 | 0.616 |
|  | Multivariable | WHO grade | **1.73** | **1.19–2.51** | **0.004** | — |
| **Benchmark** |  |  |  |  |  |  |
|  | Univariable | Chen 34-gene panel | 0.92 | 0.72–1.17 | 0.49 | 0.552 |

Total macrophage: Schoenfeld PH test global p = 0.959. Decomposed ratio: Schoenfeld PH test global p = 0.61; permutation p = 0.46 (10,000 permutations). Formal paired C-index comparison (WHO vs WHO+ratio): delta-C = +0.022, z = 1.27, p = 0.20. This analysis is limited by single-institution dominance (UW/FHCC: 90.4% of events) and should be interpreted as hypothesis-generating. The decomposed ratio results are from a companion analysis [22] using the same meta-cohort survival subset.

To test whether the null result reflected failure to distinguish prognostically distinct myeloid subpopulations, we decomposed the myeloid compartment into microglia-like and macrophage-like fractions using ssGSEA gene sets validated against single-cell ground truth (pseudo-bulk Spearman r = 0.77, 95% CI 0.54–0.89, p = 6.6 × 10^−6^; conservative estimate removing gene set overlap: r = 0.70, p = 0.0001) [22]. Despite this biological validity, the microglia-to-macrophage ratio did not predict RFS (univariable HR = 0.92, 95% CI 0.72–1.16, p = 0.46; multivariable HR = 0.98, 95% CI 0.77–1.24, p = 0.85; permutation p = 0.46; Table 3). Neither total macrophage burden nor decomposed myeloid composition improved upon WHO grade alone (C-index: WHO = 0.594; WHO + macro_score = 0.576; WHO + ratio = 0.616; formal paired comparison: delta-C = +0.022, z = 1.27, p = 0.20) [22].

As a benchmark, the Chen et al. 34-gene expression panel [5], validated at AUC 0.81 in 1,856 patients, also achieved near-chance discrimination (C-index = 0.552, HR = 0.92, p = 0.49) [22], suggesting cohort-specific limitations (small sample, single-institution dominance at 90.4% of events from UW/FHCC) rather than a TME-specific failure.

An exploratory NF2 expression proxy analysis revealed a suggestive differential effect: in the NF2-low subset (enriched for NF2-mutant tumors; N = 51, 36 events), the microglia/macrophage ratio showed a near-significant trend (HR = 0.68, 95% CI 0.46–1.01, p = 0.056), while in NF2-high tumors (N = 50, 37 events) the ratio was null (HR = 0.98, p = 0.89) [22], consistent with the Maas framework predicting NF2-specific prognostic relevance [16].

All models satisfied proportional hazards assumptions (Schoenfeld global p = 0.959 for total macrophage; p = 0.61 for ratio). CIBERSORTx-derived macrophage tertile yielded concordant null results in sensitivity analysis (continuous HR = 1.147, p = 0.721; high vs low HR = 0.987, p = 0.963). T-cell score and CSF1R pathway score were also non-prognostic (Supplementary Table S4).

To directly benchmark our bulk-deconvolution myeloid score against the Maas 2026 single-cell-anchored framework, we computed single-sample enrichment scores for published Maas microglia-like markers (TMEM119, P2RY13, P2RY12, GPR34, SLC2A5, CX3CR1, SALL1, OLFML3) and peripheral-macrophage markers (C3, F10, EMILIN2, F5, GDA, SELL, HP, VCAN, S100A8, S100A9) across the harmonized cohort (N = 968). Across the full cohort, both component macrophage scores correlated strongly with the Maas microglia-like signature (EPIC Macrophages Spearman ρ = 0.69; MCPcounter

Monocytic lineage ρ = 0.71; CIBERSORTx absolute macrophage score ρ = 0.58; all p < 10^−80^) but only modestly with the Maas peripheral-macrophage signature (EPIC ρ = 0.35; MCPcounter ρ = 0.18; CIBERSORTx ρ = 0.14; all p < 2.4 × 10^−5^). On the cluster-linked subset (N = 234), the z-scored consensus score replicated this asymmetry (microglia-like ρ = 0.73; peripheral-macrophage ρ = 0.24, p = 2.2 × 10^−4^). This quantifies a specific limitation of bulk RNA-seq deconvolution in meningioma: the macrophage consensus score is dominated by the brain-resident microglia-like compartment and captures only a weak signal from peripheral-BMDM infiltration — the population whose expansion drives the NF2-adjusted Maas risk continuum. Neither the microglia-like nor peripheral-macrophage ssGSEA scores predicted RFS (all p ≥ 0.13), consistent with the Maas observation that it is the qualitative microglia-to-BMDM shift, rather than total myeloid abundance, that carries prognostic information. Cluster B was the highest-scoring subtype for both microglia-like (Kruskal–Wallis p = 5.4 × 10^−13^) and peripheral-macrophage (p = 1.9 × 10^−4^) signatures, with the microglia-to-peripheral ratio also maximal in Cluster B (p = 2.2 × 10^−7^), reinforcing the subtype-specific TAM enrichment reported above.

Notably, this survival analysis is explicitly exploratory. The UW/FHCC dataset contributed 69/102 samples (67.6%) and 66/73 events (90.4%), and 315-month censored observations were excluded per protocol, generating a selection bias toward high-event-rate samples (95.7% event rate in the UW/FHCC subset). An adequately powered multi-institutional survival analysis with balanced censoring across contributing datasets is needed to definitively test immune-prognostic associations in meningioma.

## DISCUSSION

### A Novel Estrogen–Immune Dissociation in Meningioma

The central finding of this study is a previously unreported dissociation between maintained ESR1 expression and suppressed estrogen-responsive transcriptional programs in macrophage-infiltrated meningiomas, confirmed across a multi-layer sensitivity battery (eleven analyses including reference-matrix-independent, purity-adjusted, rotation-based self-contained, and empirical-null tests). The individual receptor analysis sharpens this observation: ESR1 is upregulated (log_2_FC = +0.40) while the functionally dominant hormone receptor in meningioma PGR is downregulated (log_2_FC = −0.34), and ESR2 is unchanged. This pattern indicates post-receptor signaling disruption rather than receptor silencing.

The within-cluster analyses provide important mechanistic context. The estrogen–immune dissociation persists significantly within both Cluster A (FDR = 2.8 × 10^−3^ / 3.6 × 10^−3^, N = 82) and Cluster B (FDR = 7.0 × 10^−3^ / 1.9 × 10^−2^, N = 39), with a non-significant cluster × macrophage interaction (FDR = 0.93 / 0.98), indicating a cross-subtype TAM-associated phenomenon rather than a finding confined to a single transcriptomic context. This argues against NF2-loss-specific biology as the sole explanation and suggests a more general mechanism by which myeloid infiltration remodels hormone-responsive transcription. The robustness battery with self-contained fry test (FDR ≤ 1.4 × 10^−12^), empirical random-gene-set null (rank 1/1,000), and meningothelial-lineage purity proxy (FDR = 1.6 × 10^−6^) further argues against compositional dilution, inter-gene-correlation assumptions, or purity-macrophage collinearity as the signal source.

A key interpretive caveat is that bulk RNA-seq cannot attribute the observed ESR1 upregulation to tumor cells versus infiltrating macrophages. Macrophages are known to express ESR1 [12], and the observed ESR1 increase in macrophage-high tumors could partly reflect macrophage-contributed transcripts rather than tumor-cell receptor upregulation. Similarly, the suppression of estrogen response gene sets could reflect compositional dilution rather than active transcriptional disruption, with the reduced tumor cell fraction in macrophage-infiltrated samples contributing fewer tumor-derived transcripts. The purity-adjusted analysis (EPIC otherCells covariate: FDR = 7.3 × 10^−5^; meningothelial-lineage proxy: FDR = 1.6 × 10^−6^) addresses this partially, but otherCells is a composite score that does not specifically control for macrophage-contributed ESR1. Single-cell RNA-seq is required to determine which cell populations carry the estrogen response signal and whether the dissociation reflects tumor-cell biology, macrophage biology, or both. With this caveat, several non-mutually exclusive mechanisms may underlie this dissociation. First, inflammatory mediators enriched in macrophage-high tumors, particularly IL-6/JAK/STAT3 and IFN-γ pathways, both significantly upregulated in our data, may disrupt downstream estrogen-responsive transcription while leaving ESR1 expression intact or even inducing it as a compensatory response. IL-6/STAT3 signaling is known to antagonize estrogen receptor transcriptional activity in other systems [18], and the concurrent upregulation of both inflammatory and ESR1 expression is consistent with this model. Second, PGR downregulation may reflect subtype-intrinsic biology: NF2-loss Cluster B tumors (the macrophage-enriched subtype) may inherently express lower PGR as part of their molecular identity, partially independent of immune composition. Third, the Hallmark ESTROGEN_RESPONSE gene sets contain proliferative and metabolic genes beyond classical receptor targets; their suppression in immune-infiltrated tumors may reflect a broader transcriptional reprogramming in the context of a myeloid-dominant microenvironment, supported by concurrent downregulation of MYOGENESIS and SPERMATOGENESIS gene sets.

This finding is distinct from immune–endocrine interactions reported in other cancers. In triple-negative breast cancer, TAMs suppress ERβ expression via PI3K/AKT/FOXO3a [32], a receptor-silencing mechanism. In endometrial cancer, macrophage-derived IL-17A upregulates ERα through epigenetic mechanisms [20]. Our observation of simultaneous ESR1 *upregulation* and estrogen-responsive program *suppression* constitutes a third pattern: post-receptor transcriptional disruption in the context of maintained — and even increased — receptor expression. This pattern has not been described in meningioma or in other CNS tumors.

The clinical implication is that macrophage-infiltrated meningiomas may lose hormonal responsiveness despite retaining estrogen receptor expression. If confirmed at the protein level, this dissociation would have direct relevance for hormonal therapy trials: progestational therapies (mifepristone and others) may be most effective in immune-desert subtypes where the estrogen-responsive transcriptional program is intact, while macrophage-infiltrated tumors may require combined hormonal and immune-targeting strategies. Future trials evaluating hormonal agents in meningioma should consider stratification by immune phenotype.

These findings intersect with a long-standing puzzle in meningioma hormonal biology. Progesterone receptor positivity is present in ∼70% of WHO grade I tumors and has traditionally been treated as a favorable prognostic marker [6]; conversely, the 2:1 female-to-male incidence ratio, pregnancy-associated growth acceleration, and epidemiologic associations between long-term progestin exposure (cyproterone acetate, chlormadinone acetate, nomegestrol acetate) and meningioma onset [4, 21] all indicate biologically consequential hormonal signaling. Yet clinical trials of anti-progestational therapy (most notably mifepristone) have yielded inconsistent results, and no hormonal agent has gained regulatory approval for meningioma [8]. One parsimonious interpretation of that history, in light of the present findings, is that hormonal responsiveness is subtype-and TME-dependent: tumors with intact estrogen-responsive transcriptional programs may respond, while macrophage-infiltrated tumors with maintained ESR1 but suppressed downstream programs may be pharmacologically refractory. This predicts that PGR immunohistochemistry combined with macrophage quantification (e.g., CD68 or PU.1 IHC) could prospectively stratify candidates for anti-progestin or selective estrogen receptor modulator trials, identifying a hormonally competent, macrophage-low population most likely to benefit.

The sex-adjustment result supports the biological interpretation. The persistence of a significant association after sex adjustment (FDR = 1.9 × 10^−6^, N = 79) is inconsistent with sex as the sole driver, though the small subsample size (12% of the tertile comparison population, predominantly from UW/FHCC) limits definitive conclusions about signal amplification. The convergence of eleven complementary sensitivity analyses, each using a different analytical specification, provides robust triangulation despite the limitations of any individual analysis.

### Subtype-Dependent Myeloid Architecture and CSF1R Targeting

The organization of myeloid infiltration along transcriptomic subtype boundaries rather than WHO grade (Kruskal–Wallis p = 7.4 × 10^−16^ vs p = 0.399) has direct translational implications. Cluster B, characterized by NF2 loss [30], exhibited the highest macrophage fraction by three-method consensus and the highest CSF1R and CSF1 transcript abundance, consistent with a TAM-rich microenvironment sustained by paracrine CSF1–CSF1R signaling. CSF1R ranked among the top differentially expressed genes in macrophage-high tumors regardless of deconvolution reference (rank 29 by MCPcounter-based DE; rank 13 by CIBERSORTx-based DE), confirming this is not a reference-matrix artifact.

An important question is the cellular source of CSF1R expression. While the concordance between macrophage infiltration and CSF1R transcript abundance is consistent with macrophage-expressed CSF1R sustaining autocrine/paracrine TAM survival, bulk RNA-seq cannot exclude tumor-intrinsic CSF1R expression. Guo et al. identified CSF1R transcripts in meningioma tumor cells at single-cell resolution [9], and Lan et al. demonstrated that SYHA1813 directly inhibits meningioma cell proliferation via P53 pathway activation [13], suggesting tumor-intrinsic CSF1R signaling independent of macrophage biology. Both interpretations support CSF1R inhibitor deployment in Cluster B. Notably, Cluster B is molecularly characterized by NF2 loss [30], and the subtype-dependent myeloid architecture may be substantially NF2-driven. Teranishi et al. showed that NF2-associated meningiomas have lower tumor purity (74.4% vs 80.9%, P = 0.02), higher M2 macrophage fractions (P = 0.005), and elevated CD68+ density (P = 0.007) [28], consistent with NF2 loss promoting macrophage recruitment through YAP/TAZ-mediated chemokine secretion.

The rationale for CSF1R-targeted therapy over checkpoint blockade is reinforced by clinical data: pembrolizumab achieved 0% objective response rate despite PD-L1 positivity in 88% of Grade II–III meningiomas [2], indicating that the immunosuppressive TAM-dominated TME, rather than absent PD-L1 engagement, is the primary barrier. The dual VEGFR/CSF1R inhibitor SYHA1813 has shown Phase I activity in recurrent high-grade gliomas (ORR 18.8%) with confirmed CNS penetration [11], and a Phase II trial in recurrent meningioma is registered (NCT06739213). This CSF1R candidate-biomarker rationale is independently endorsed by Lötsch et al. [15] and Lan et al. [13], who demonstrated direct antitumor activity of SYHA1813 in meningioma xenografts via P53 pathway activation, although the pooled signal from early-phase trials of CSF1R inhibitors in CNS tumors remains modest, warranting subtype-enriched rather than unselected enrollment. Clinical trials should stratify patients by transcriptomic subtype to detect subtype-specific activity — Cluster B patients represent the biologically optimal candidates, while immune-desert subtypes (Cluster C) may require alternative strategies.

### Bulk RNA-seq Cannot Prognosticate Myeloid Biology in Meningioma

Both total macrophage burden (this study: HR = 0.904, p = 0.53) and the decomposed microglia-to-macrophage ratio (HR = 0.92, p = 0.46) [22] failed to predict recurrence-free survival, despite the ratio’s biological validity (pseudo-bulk r = 0.77 vs single-cell ground truth). The progression from coarse (total macrophage) to refined (microglia/macrophage composition) myeloid quantification with equivalent null results argues against analytical imprecision as the sole explanation.

This double-negative finding contrasts with positive IHC-based results: Lötsch et al. reported HR = 2.11 (p = 0.023) for CD68-positive TAM density [15], and Maas et al. demonstrated HR = 2.00 (p = 2.4 × 10^−5^) for PU.1-based TAM quantification [16]. The common thread between these positive results and our negative ones is the measurement modality. IHC captures spatial abundance of specific cell populations at protein level, while bulk RNA-seq deconvolution estimates fractional composition from a mixed transcriptome, losing spatial distribution, polarization state, and protein-level information that may carry the prognostic signal. Our Maas-proxy head-to-head (ρ = 0.73 with microglia-like markers vs ρ = 0.24 with peripheral-macrophage markers) directly quantifies this: the consensus bulk-RNA-seq macrophage score captures predominantly the brain-resident microglia compartment and only weakly registers the peripheral-BMDM expansion that drives the Maas NF2-adjusted risk continuum. The null RFS association in our data is therefore biologically consistent with Maas 2026 rather than contradictory.

Several cohort-specific factors compound this modality limitation. First, the Maas risk continuum is defined within NF2-mutant meningiomas [16]; our unselected cohort includes an estimated 30–40% NF2-wild-type tumors, diluting any NF2-specific prognostic signal. Accounting for proxy measurement error (the ssGSEA ratio captures ∼22% of MC risk class variance) and NF2-wild-type dilution, the Maas IHC HR of 2.00 would attenuate to approximately 1.24, for which our study had approximately 15% power [22]. The exploratory NF2-low trend (HR = 0.68, p = 0.056) supports this interpretation. A quantitative attenuation analysis estimates that the Maas IHC HR of 2.00 would attenuate to approximately 1.24 after proxy measurement error and NF2-wild-type dilution, yielding only 15% power with 73 events [22].

Second, the survival cohort is small (N = 101–102, 73 events) and dominated by a single institution (UW/FHCC: 90.4% of events). Third, the Chen et al. 34-gene panel, validated at AUC 0.81 in 1,856 patients, also achieved near-chance discrimination (C-index = 0.552), confirming that the null finding reflects cohort-level limitations rather than a TME-specific failure.

These findings establish a methodological boundary: bulk RNA-seq deconvolution characterizes TME composition but cannot prognosticate it in moderately sized, NF2-unselected cohorts. Clinical implementation of TME-based risk stratification will require either protein-level assays, methylation-based deconvolution, or substantially larger bulk RNA-seq cohorts with confirmed NF2 status.

### Limitations

Overall, this is a hypothesis-generating bulk-RNA-seq computational study: the estrogen–immune dissociation is a robust statistical association across eleven sensitivity analyses but remains an association requiring single-cell and functional validation, and the survival null is underpowered against the realistic attenuated effect size (∼HR 1.24) expected after accounting for proxy measurement error and NF2-wildtype dilution. Several limitations warrant acknowledgment. First, this is a computational study using publicly available data; no tissue-level validation (IHC, single-cell RNA-seq) was performed. The estrogen–immune dissociation is associational and requires mechanistic validation to determine which cell populations carry the estrogen response signal and whether TAM-secreted inflammatory mediators directly suppress estrogen-responsive transcription in meningioma cells. Second, bulk RNA-seq deconvolution using the LM22 reference cannot distinguish meningioma-specific border-associated macrophages from conventional macrophages; a meningioma-specific reference matrix would improve resolution. Third, Thirimanne cluster linkage required probabilistic matching for Baylor (N = 18) and UCSF 2022 (N = 15) samples, and the HKU/UCSF dataset (N = 302) could not be linked due to absence of a public identifier crosswalk. Fourth, WHO grade annotation reflects multiple classification eras (WHO 2000–2021), and NF2 mutation status could not be directly assessed without matched genomic data.

Fifth, survival analysis was exploratory and institution-imbalanced; the minimum detectable HR at 80% power is approximately 1.40, meaning true moderate effects cannot be excluded. Sixth, the microglia/macrophage ratio survival results derive from the same meta-cohort survival subset [22], limiting independence from the total macrophage analysis. Seventh, the sex-adjusted camera analysis was performed on a convenience subsample of 79 sex-annotated patients; although the signal strengthened, this represents 12% of the tertile comparison population.

## CONCLUSIONS

This study reveals a previously unreported dissociation between maintained ESR1 expression and suppressed estrogen-responsive transcriptional programs in macrophage-infiltrated meningiomas, confirmed across a multi-layer sensitivity battery (eleven analyses including reference-matrix-independent, purity-adjusted, rotation-based self-contained, and empirical-null tests) and robust to sex and tumor purity confounding. The finding connects two established axes of meningioma biology (hormonal dependence and myeloid microenvironment) and has direct implications for hormonal therapy stratification. Myeloid architecture is organized by transcriptomic subtype rather than WHO grade, with the NF2-associated Cluster B exhibiting concordant enrichment of macrophages, CSF1R, and CSF1, identifying it as the candidate subtype for subtype-selective enrollment in CSF1R-directed clinical trials. Neither total macrophage score nor the decomposed microglia-to-macrophage ratio predicted recurrence-free survival in bulk RNA-seq, establishing that this modality detects but cannot prognosticate TME composition. Single-cell validation of the estrogen–immune interface and immune phenotype stratification in hormonal therapy trials are the critical next steps.

## List of Abbreviations

BAM: Border-associated macrophage
CAF: Cancer-associated fibroblast
CSF1R: Colony-stimulating factor 1 receptor
DE: Differential expression
EPIC: Estimating the Proportions of Immune and Cancer cells
EPV: Events per variable
FDR: False discovery rate
GEO: Gene Expression Omnibus
HR: Hazard ratio
IHC: Immunohistochemistry
KW: Kruskal–Wallis
LASSO: Least absolute shrinkage and selection operator
LM22: 22-cell leukocyte reference matrix (CIBERSORTx)
MCP: Microenvironment Cell Populations counter
PH: Proportional hazards
PGR: Progesterone receptor
RFS: Recurrence-free survival
ssGSEA: Single-sample gene set enrichment analysis
TAM: Tumor-associated macrophage
TME: Tumor microenvironment
TPM: Transcripts per million
WHO: World Health Organization

## DECLARATIONS

### Ethics approval

Not applicable. All data were obtained from publicly available, de-identified datasets deposited in the Gene Expression Omnibus. Informed consent and IRB approval are not required for analysis of publicly available de-identified data.

### Consent to participate

Not applicable.

### Consent for publication

Not applicable.

### Availability of data and materials

All expression data are publicly available from GEO under accession numbers GSE212666, GSE252291, GSE183653, GSE136661, and GSE101638. The Maas snRNA-seq Seurat object is available from Zenodo (doi:10.5281/zenodo.17296825). Analysis code and the intermediate data objects required to reproduce all reported numbers are available at a private GitHub repository (access on request during peer review; public release on acceptance).

### Code availability

Analysis code and the intermediate data objects required to reproduce all reported numbers are available at a private GitHub repository (access on request during peer review; public release on acceptance).

### Author contributions

DP conceived the study, performed all computational analyses, and drafted the manuscript. All authors read and approved the final manuscript.

## Acknowledgements

The authors thank the groups of Thirimanne, Holland, Raleigh, Choudhury, and Maas for making their meningioma transcriptomic and single-cell datasets publicly available, and Stanford University for access to the CIBERSORTx computational platform.

## GRAPHICAL ABSTRACT

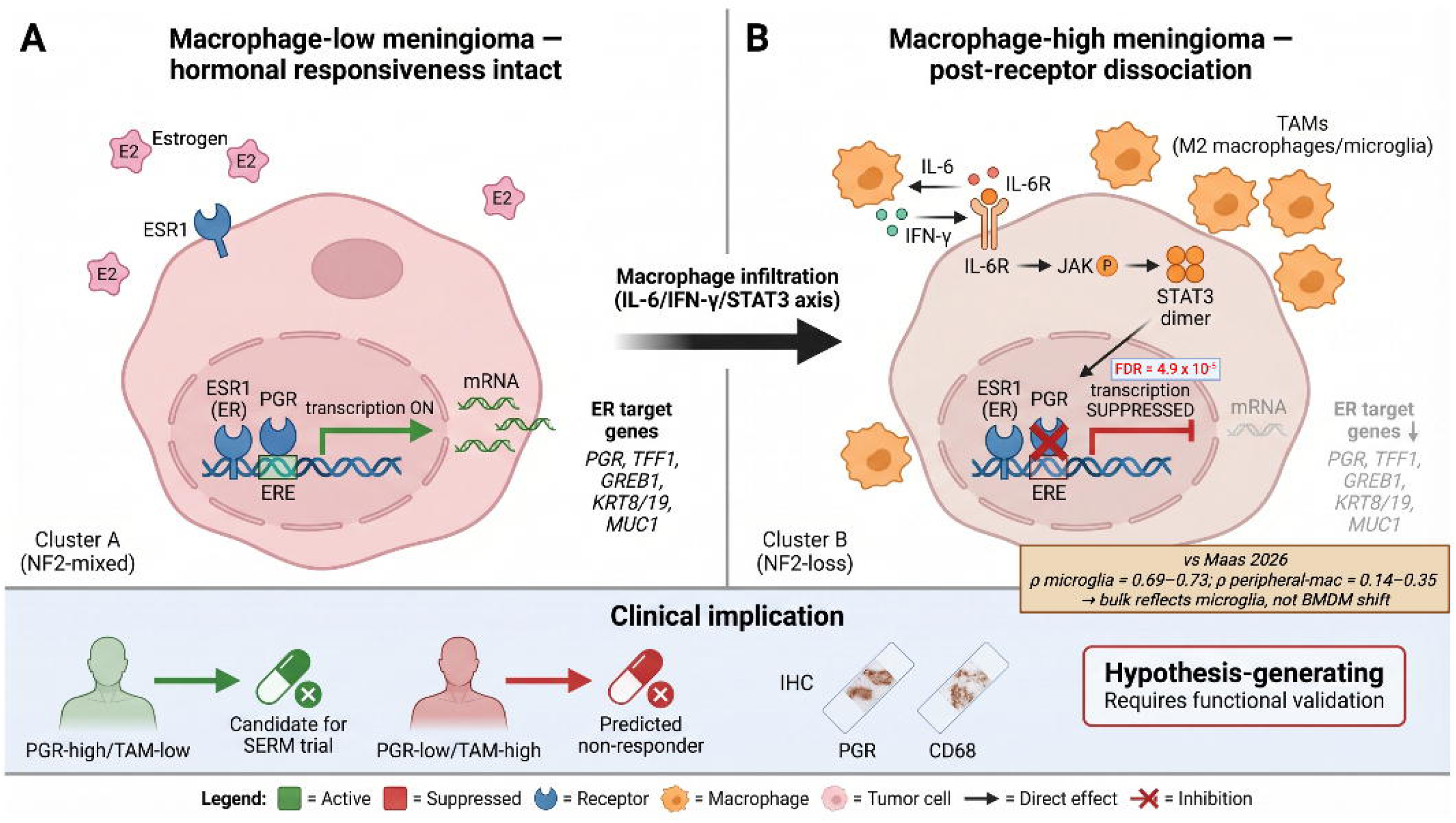

**Conceptual model of the estrogen–immune dissociation in macrophage-infiltrated meningioma.** Two-state schematic contrasting hormone-responsive (macrophage-low, Cluster A NF2-mixed) and dissociated (macrophage-high, Cluster B NF2-loss) tumor states. In the macrophage-high state, TAM-derived IL-6 and IFN-γ engage IL-6R/JAK/STAT3 signaling that disrupts downstream estrogen-responsive transcription despite maintained — or modestly increased — ESR1 expression; PGR is concomitantly downregulated. The Maas concordance callout summarizes the head-to-head correlation of our consensus macrophage score with Maas 2026 microglia-like markers (ρ = 0.69–0.73) versus peripheral-macrophage markers (ρ = 0.14–0.35), indicating that the bulk RNA-seq signal predominantly reflects the brain-resident microglia compartment rather than the peripheral-BMDM expansion that drives the Maas methylome-based risk continuum. The bottom banner illustrates the candidate prospective stratification (PGR + CD68/PU.1 IHC) for future anti-progestin and SERM trials, framed as hypothesis-generating pending functional validation. Created with BioRender.com.

## SUPPLEMENTARY MATERIAL

### SUPPLEMENTARY METHODS

**Batch correction quality control.** Of 17,585 × 968 values, 1,277,357 (7.5%) were clipped from negative to zero after limma::removeBatchEffect, consistent with expected behavior of the shift-and-mean method. The primary endpoint cell type (EPIC macrophages) exhibited a fully continuous distribution with no samples at the zero floor (range: 0.000–0.248; Q1 = 0.025, Q3 = 0.064). PCA before and after batch correction, colored by batch identity and WHO grade independently, is shown in Supplementary Fig. S1. **Crosswalk methodology.** Thirimanne cluster linkage strategy details: (1) direct sample-ID matching for UW/FHCC (N = 201 linked); (2) probabilistic matching using age ±1 year, biological sex, and WHO grade for Baylor (N = 18); (3) M-number and age ±2 year matching for UCSF 2022 (N = 15). HKU/UCSF lacked a public MNG-to-QM crosswalk and was not linked.

**CIBERSORTx p < 0.05 subset concordance.** In the 279 samples reaching CIBERSORTx significance, macrophage concordance remained robust across 2/3 method pairs (EPIC × MCPcounter r = 0.639; MCPcounter × CIBERSORTx r = 0.640; EPIC × CIBERSORTx r = 0.434). A two-method composite (EPIC + MCPcounter) yielded identical cluster rankings (Spearman r = 1.00 vs the 3-method composite).

---

**Supplementary Table S1.** Therapeutic target expression across Thirimanne transcriptomic subtypes (A–G).

|  | Cluster A | Cluster B | Cluster C | Cluster D | Cluster E | Cluster F | Cluster | KW p- |
| --- | --- | --- | --- | --- | --- | --- | --- | --- |
| Gene | (N=82) | (N=39) | (N=46) | (N=37) | (N=11) | (N=8) | G (N=2) | value |
| CSF1R | 5.11 | **6.60** | 4.71 | 4.82 | 3.84 | 4.87 | 5.21 | $1.4 \times 10^{-14}$ |
| CSF1 | 5.19 | **6.31** | 4.53 | 5.14 | 3.80 | 4.41 | 4.09 | $9.4 \times 10^{-17}$ |
| IL34 | 3.98 | 4.43 | 4.40 | **4.84** | 4.78 | 2.89 | 2.10 | <0.001 |
| CD274 | 0.62 | 0.60 | 0.87 | 0.82 | 0.78 | **1.77** | 1.03 | <0.001 |
| CDK4 | 6.23 | 6.21 | 5.81 | 5.82 | 5.47 | 6.06 | **6.52** | <0.001 |
| CDK6 | 2.52 | 2.57 | 1.92 | 1.46 | 1.48 | **4.17** | 3.11 | $1.8 \times 10^{-17}$ |
| VEGFA | 3.97 | 2.83 | 4.13 | 4.97 | 5.81 | **7.26** | 5.12 | <0.001 |
| PDGFRA | 1.54 | 1.84 | 0.81 | 1.05 | 0.41 | 0.63 | **2.51** | <0.001 |

Values are median log-CPM. **Bold** = highest cluster. N = 225 cluster-linked samples.

**Supplementary Table S2.**
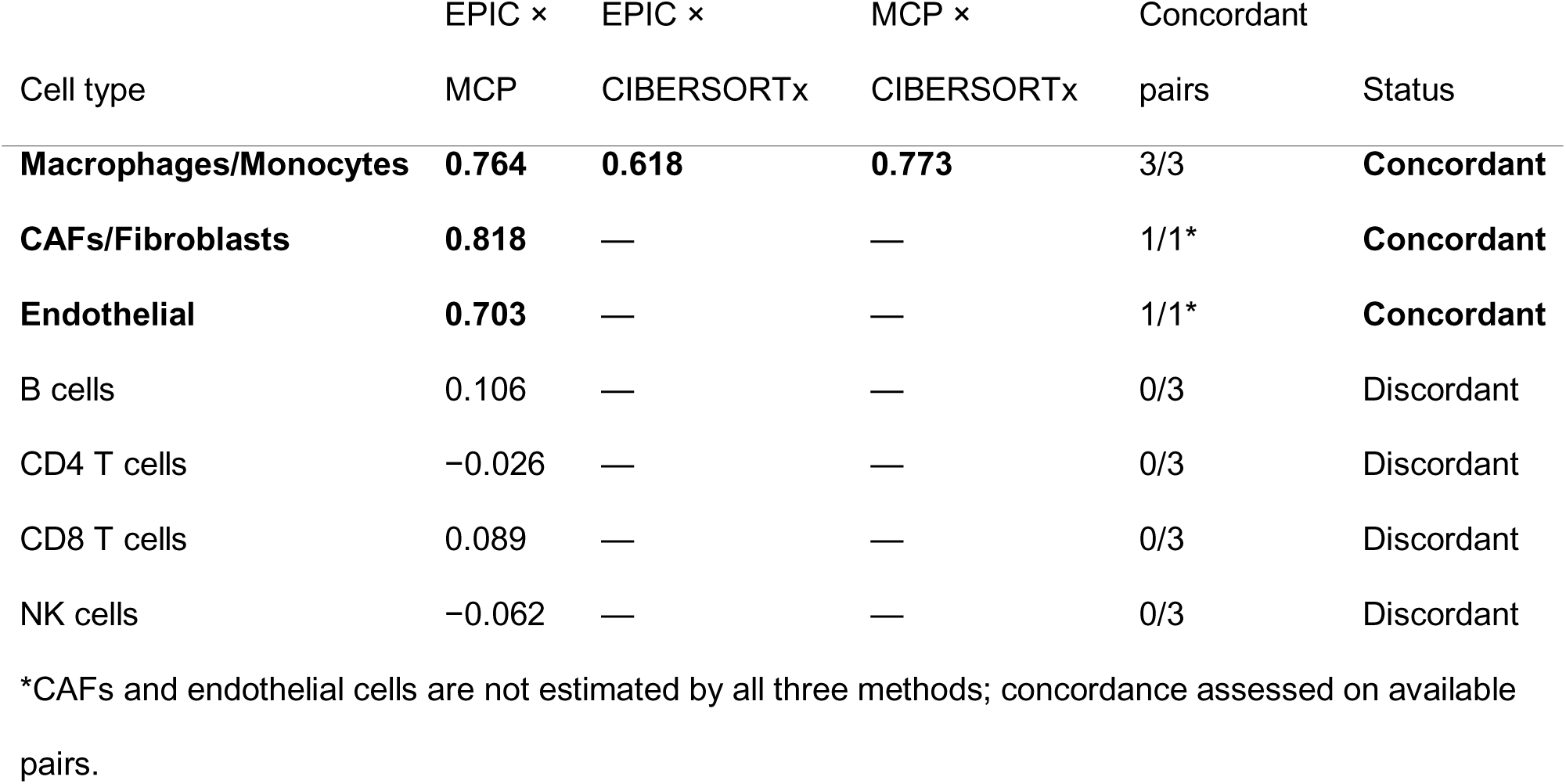
Cross-method immune deconvolution concordance matrix.

| Cell type | EPIC × | EPIC × | MCP × | Concordant |  |
| --- | --- | --- | --- | --- | --- |
|  | MCP | CIBERSORTx | CIBERSORTx | pairs | Status |
| **Macrophages/Monocytes** | **0.764** | **0.618** | **0.773** | 3/3 | **Concordant** |
| **CAFs/Fibroblasts** | **0.818** | — | — | 1/1* | **Concordant** |
| **Endothelial** | **0.703** | — | — | 1/1* | **Concordant** |
| B cells | 0.106 | — | — | 0/3 | Discordant |
| CD4 T cells | −0.026 | — | — | 0/3 | Discordant |
| CD8 T cells | 0.089 | — | — | 0/3 | Discordant |
| NK cells | −0.062 | — | — | 0/3 | Discordant |

\*CAFs and endothelial cells are not estimated by all three methods; concordance assessed on available pairs.

**Supplementary Table S3.** Dunn post-hoc pairwise comparisons for macrophage score across clusters.

| Comparison | Z statistic | Adjusted p (BH) | Significant |
| --- | --- | --- | --- |
| A – B | -4.507 | $1.31\times10^{-5}$ | Yes |
| A – C | 4.616 | $9.78\times10^{-6}$ | Yes |
| A – D | 2.506 | 0.017 | Yes |
| A – E | 2.965 | 0.005 | Yes |
| B – C | 7.934 | $2.12\times10^{-14}$ | Yes |
| B – D | 5.983 | $1.10\times10^{-8}$ | Yes |
| B – E | 5.357 | $2.83\times10^{-7}$ | Yes |
| C – D | -1.603 | 0.136 | No |
| C – E | 0.303 | 0.762 | No |
| D – E | 1.327 | 0.205 | No |

**Supplementary Table S4.** Camera gene set enrichment results — all Hallmark gene sets with FDR <.

| Gene set | Direction | FDR | NGenes |
| --- | --- | --- | --- |
| HALLMARK_ALLOGRAFT_REJECTION | Up | $1.76 \times 10^{-12}$ | 200 |
| HALLMARK_INTERFERON_GAMMA_RESPONSE | Up | $8.76 \times 10^{-8}$ | 200 |
| HALLMARK_COMPLEMENT | Up | $2.43 \times 10^{-6}$ | 200 |
| HALLMARK_INTERFERON_ALPHA_RESPONSE | Up | $5.24 \times 10^{-6}$ | 97 |
| HALLMARK_IL6_JAK_STAT3_SIGNALING | Up | $5.24 \times 10^{-6}$ | 87 |
| HALLMARK_INFLAMMATORY_RESPONSE | Up | $1.46 \times 10^{-5}$ | 200 |
| HALLMARK_ESTROGEN_RESPONSE_LATE | Down | $2.79 \times 10^{-5}$ | 200 |
| HALLMARK_ESTROGEN_RESPONSE_EARLY | Down | $4.89 \times 10^{-5}$ | 200 |
| HALLMARK_SPERMATOGENESIS | Down | $1.45 \times 10^{-4}$ | 135 |
| HALLMARK_MYOGENESIS | Down | $2.47 \times 10^{-3}$ | 200 |
| HALLMARK_KRAS_SIGNALING_DN | Down | $3.82 \times 10^{-2}$ | 200 |

Camera analysis: CIBERSORTx macrophage tertile-high (N = 323) vs tertile-low (N = 323).

**Supplementary Table S5.**
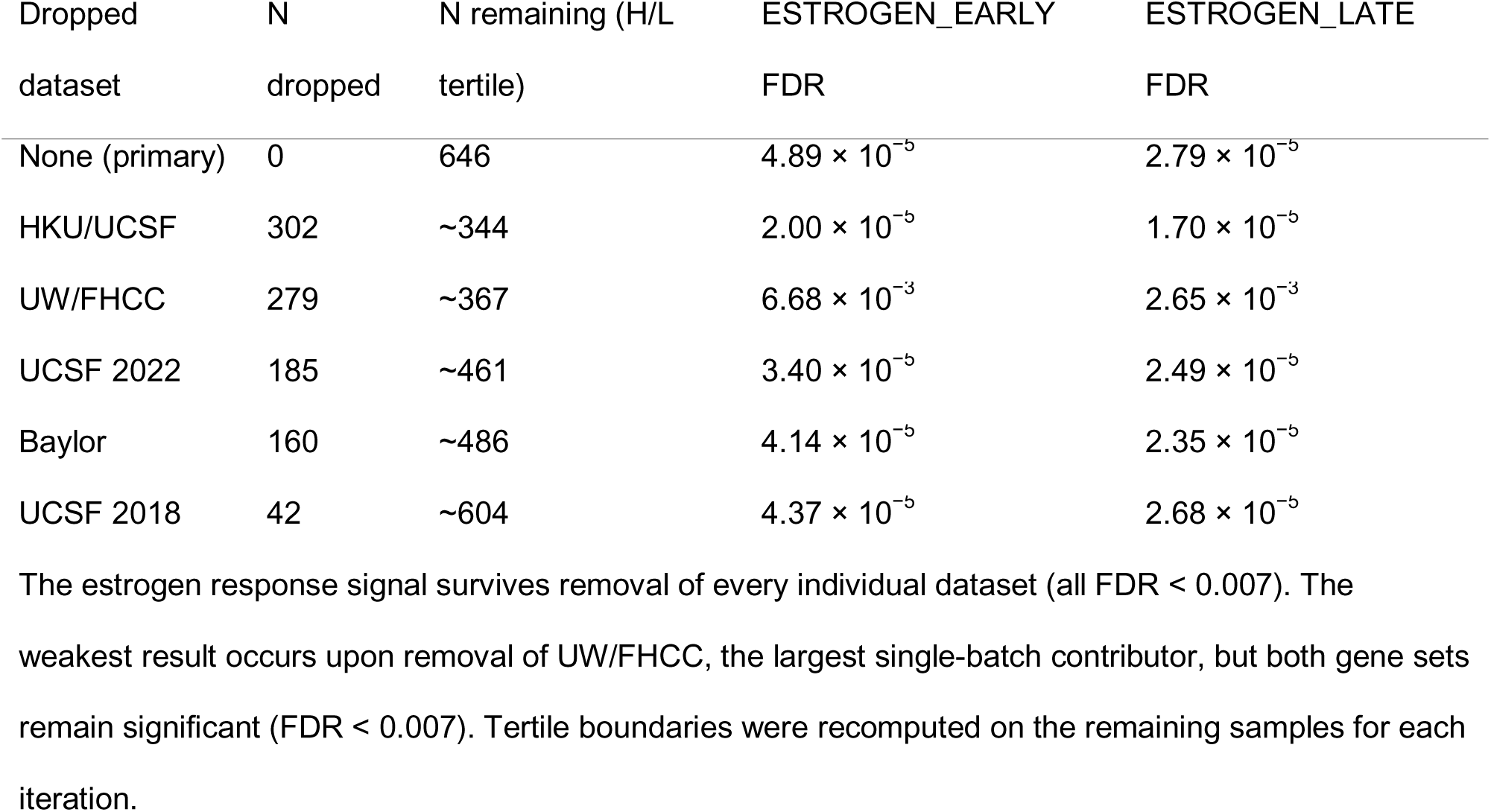
Leave-one-dataset-out camera analysis for estrogen response gene sets.

| Dropped dataset | N dropped | N remaining (H/L tertile) | ESTROGEN_EARLY FDR | ESTROGEN_LATE FDR |
| --- | --- | --- | --- | --- |
| None (primary) | 0 | 646 | $4.89 \times 10^{-5}$ | $2.79 \times 10^{-5}$ |
| HKU/UCSF | 302 | ~344 | $2.00 \times 10^{-5}$ | $1.70 \times 10^{-5}$ |
| UW/FHCC | 279 | ~367 | $6.68 \times 10^{-3}$ | $2.65 \times 10^{-3}$ |
| UCSF 2022 | 185 | ~461 | $3.40 \times 10^{-5}$ | $2.49 \times 10^{-5}$ |
| Baylor | 160 | ~486 | $4.14 \times 10^{-5}$ | $2.35 \times 10^{-5}$ |
| UCSF 2018 | 42 | ~604 | $4.37 \times 10^{-5}$ | $2.68 \times 10^{-5}$ |

The estrogen response signal survives removal of every individual dataset (all FDR < 0.007). The weakest result occurs upon removal of UW/FHCC, the largest single-batch contributor, but both gene sets remain significant (FDR < 0.007). Tertile boundaries were recomputed on the remaining samples for each iteration.

## SUPPLEMENTARY FIGURE LEGENDS

**Supplementary Fig. S1.**
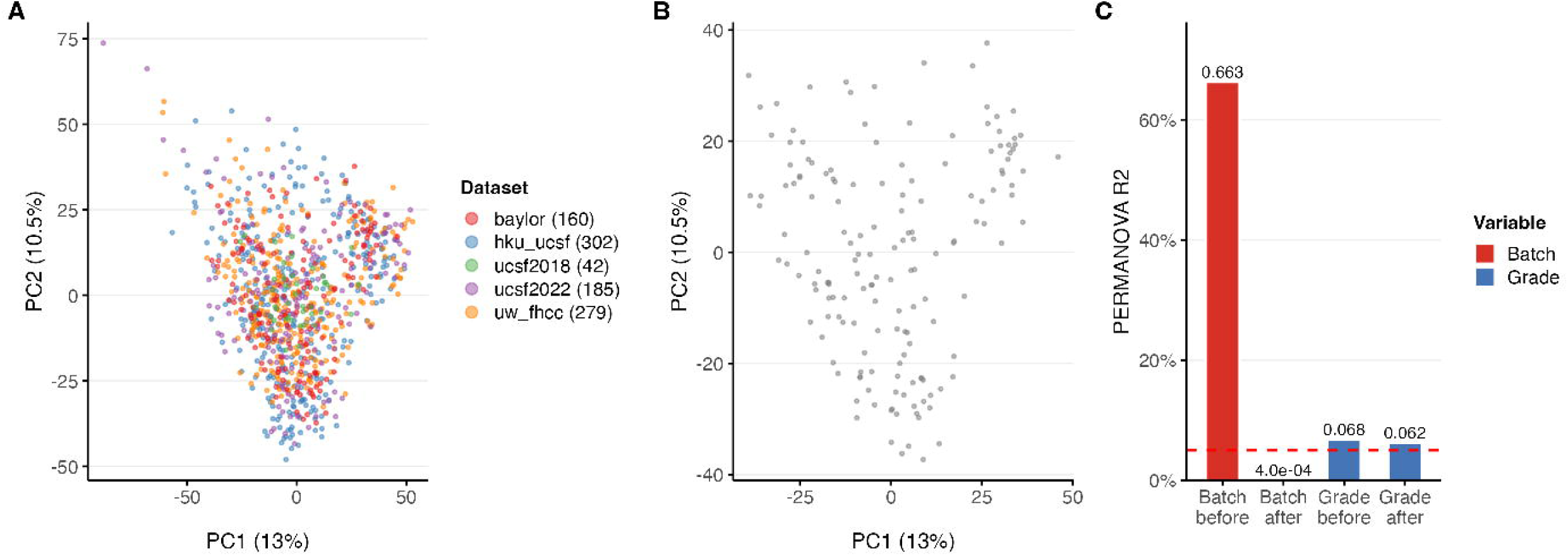
Batch correction of the 968-sample meningioma meta-cohort. (A) PCA before batch correction, colored by contributing dataset. Datasets form distinct clusters (PERMANOVA R^2^ = 0.663). (B) PCA after limma::removeBatchEffect. Dataset-explained variance reduced to R^2^ = 0.0004 while WHO grade signal preserved (R^2^ = 0.062, p = 0.002).

**Supplementary Fig. S2.**
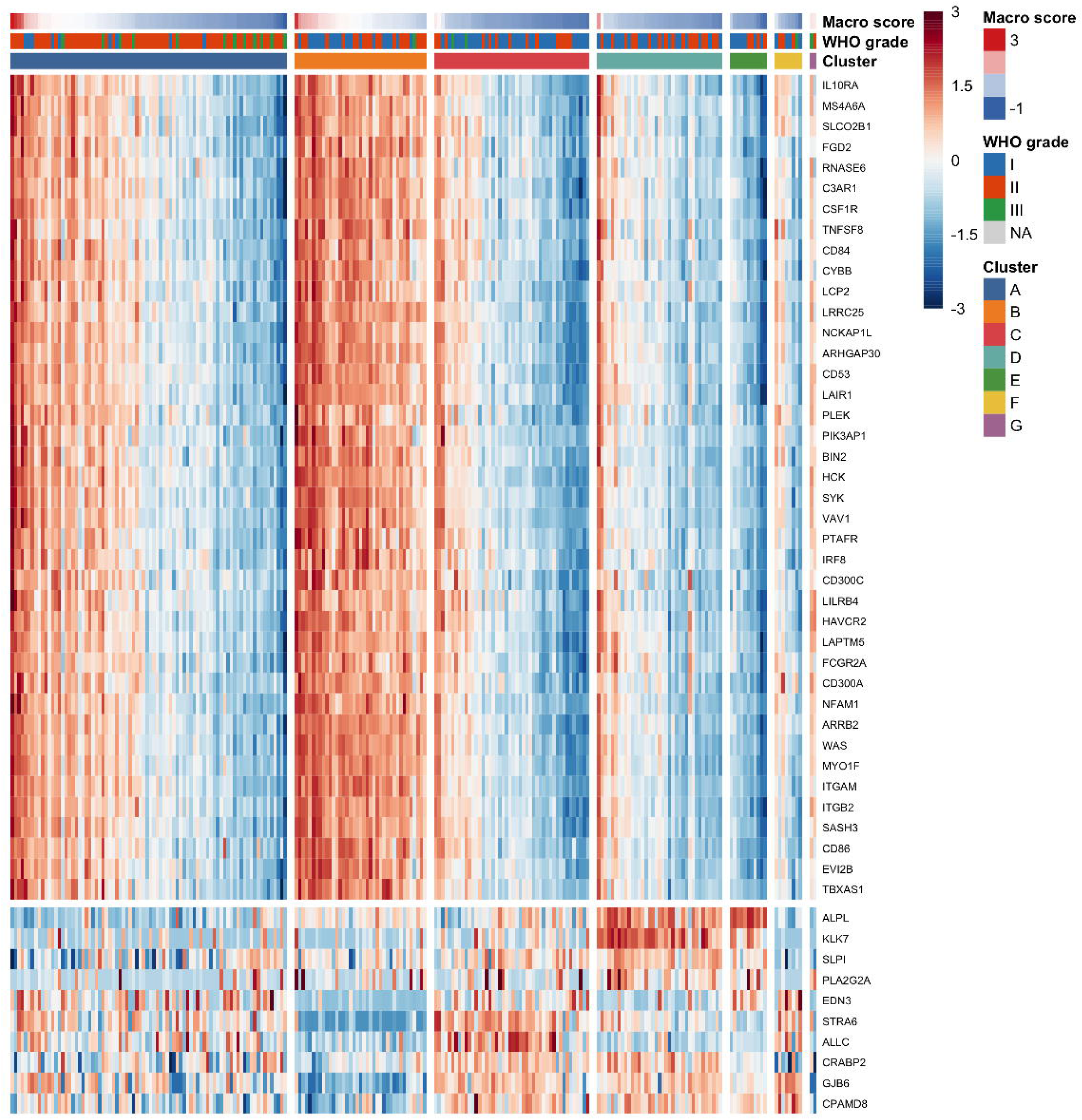
Differential expression heatmap of top 50 genes in macrophage-high versus macrophage-low tumors. Heatmap of row-z-scored expression (capped at ±3) for the top 40 upregulated and top 10 downregulated genes (by limma B-statistic) in the CIBERSORTx macrophage tertile comparison. Columns: N = 234 cluster-linked samples, ordered by cluster and macro_score within cluster. Annotations: Thirimanne cluster (A–G), WHO grade (I–III), macro_score (continuous). Ward’s D2 hierarchical clustering within up/down groups.

**Supplementary Fig. S3.**
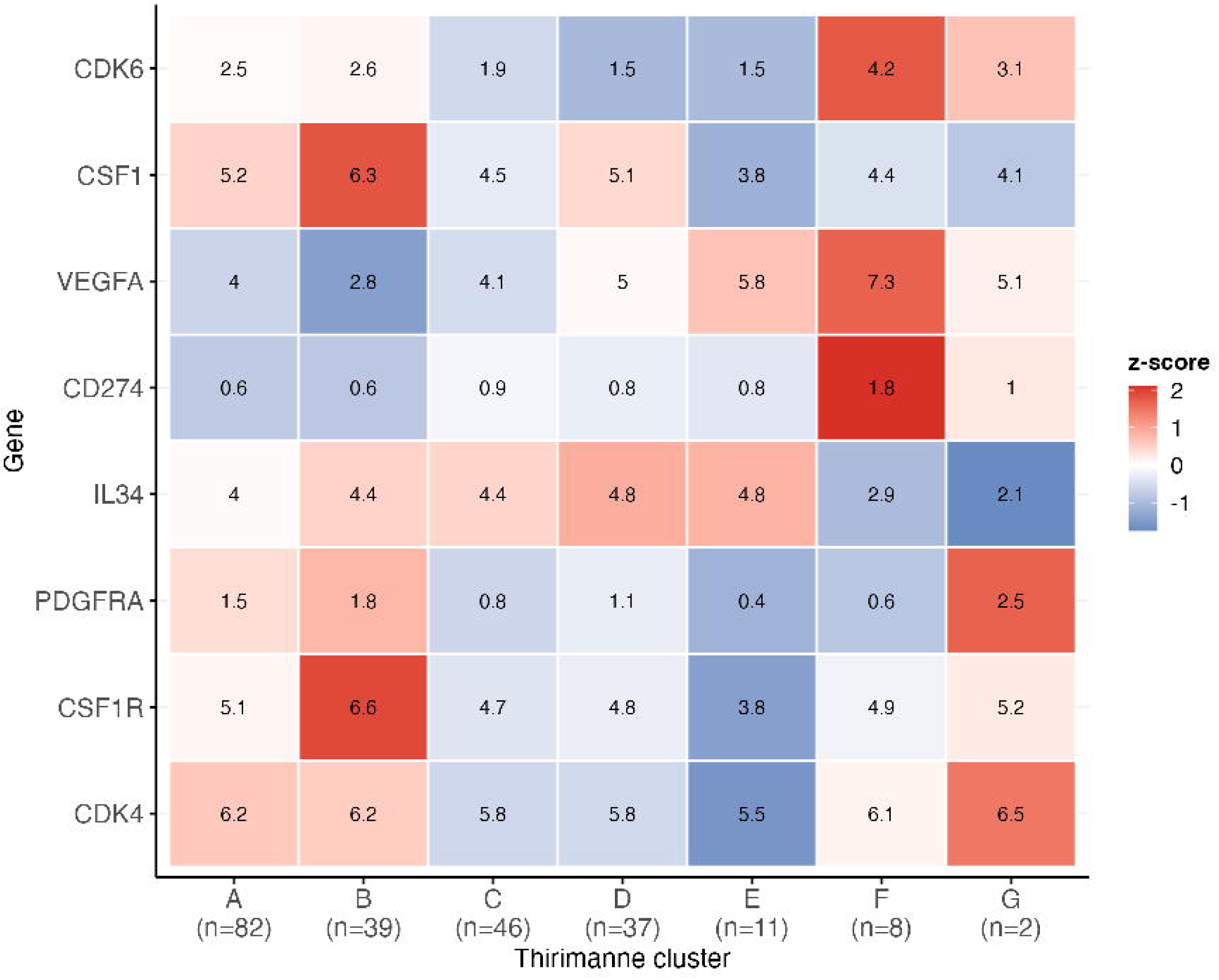
Therapeutic target expression heatmap. Heatmap of median expression (z-scored across clusters) for eight therapeutic targets across Thirimanne clusters A–G. See Supplementary Table S1 for raw values.

## REFERENCES

1. Becht E, Giraldo NA, Lacroix L, Buttard B, Elarouci N, Petitprez F, Selves J, Laurent-Puig P, Sautès-Fridman C, Fridman WH, de Reyniès A (2016) Estimating the population abundance of tissue-infiltrating immune and stromal cell populations using gene expression. Genome Biol 17:218. doi: 10.1186/s13059-016-1070-5

2. Brastianos PK, Kim AE, Giobbie-Hurder A, Lee EQ, Wang N, Eichler AF, Chukwueke U, Forst DA, Arrillaga-Romany IC, Dietrich J, Corbin Z, Moliterno J, Baehring J, White M, Lou KW, Larson J, de Sauvage MA, Evancic K, Mora J, Nayyar N, Loeffler J, Oh K, Shih HA, Curry WT, Cahill DP, Barker FG, Gerstner ER, Santagata S (2022) Phase 2 study of pembrolizumab in patients with recurrent and residual high-grade meningiomas. Nat Commun 13:1325. doi: 10.1038/s41467-022-29052-7

3. Carroll RS, Zhang J, Dashner K, Sar M, Wilson EM, Black PM (1995) Androgen receptor expression in meningiomas. J Neurosurg 82:453–460. doi: 10.3171/jns.1995.82.3.0453

4. Champeaux-Depond C, Weller J, Froelich S, Sartor A (2021) Cyproterone acetate and meningioma: a nationwide-wide population based study. J Neurooncol 151:331–338. doi: 10.1007/s11060-020-03672-9

5. Chen WC, Choudhury A, Youngblood MW, Polley M-YC, Lucas C-HG, Mirchia K, Maas SLN, Suwala AK, Won M, Bayley JC, Harmanci AS, Harmanci AO, Klisch TJ, Nguyen MP, Vasudevan HN, McCortney K, Yu TJ, Bhave V, Lam T-C, Pu JK-S, Li L-F, Leung GK-K, Chan JW, Perlow HK, Palmer JD, Haberler C, Berghoff AS, Preusser M, Nicolaides TP, Mawrin C, Agnihotri S, Resnick A, Rood BR, Chew J, Young JS, Boreta L, Braunstein SE, Schulte J, Butowski N, Santagata S, Spetzler D, Bush NAO, Villanueva-Meyer JE, Chandler JP, Solomon DA, Rogers CL, Pugh SL, Mehta MP, Sneed PK, Berger MS, Horbinski CM, McDermott MW, Perry A, Bi WL, Patel AJ, Sahm F, Magill ST, Raleigh DR (2023) Targeted gene expression profiling predicts meningioma outcomes and radiotherapy responses. Nat Med 29:3067–3076. doi: 10.1038/s41591-023-02586-z

6. Claus EB, Bondy ML, Schildkraut JM, Wiemels JL, Wrensch M, Black PM (2005) Epidemiology of intracranial meningioma. Neurosurgery 57:1088–1095; discussion 1088-1095. doi: 10.1227/01.neu.0000188281.91351.b9

7. Fan H, Li X, Zhao Y, Song L, Sun L, Wu Z, Zhang J, Zhang H, Zhang F, Huang Y, Zhang D, Wang L (2025) Decoding meningioma prognosis with multi-omics: macrophage diversity, immune-CNV interplay, and novel SPP1-targeted strategies. J Neurooncol 175:183–198. doi: 10.1007/s11060-025-05116-8

8. Grunberg SM, Weiss MH, Russell CA, Spitz IM, Ahmadi J, Sadun A, Sitruk-Ware R (2006) Long-term administration of mifepristone (RU486): clinical tolerance during extended treatment of meningioma. Cancer Invest 24:727–733. doi: 10.1080/07357900601062339

9. Guo X, Moynihan ZA, Driver J, Patel RV, Bhave VM, Maury EA, Knelson EH, Guo H, Lin J-R, Coy SM, Wang A, Gupta S, Hoffman SE, Dunn IF, Dunn GP, Petti A, Choi BD, Choudhury A, Raleigh DR, Wei K, Reardon DA, Barbie D, Lederer JA, Santagata S, Bi WL (2026) Meningioma microenvironment harbors a rich immune landscape that evolves with biological state. Neuro Oncol noag022. doi: 10.1093/neuonc/noag022

10. Hänzelmann S, Castelo R, Guinney J (2013) GSVA: gene set variation analysis for microarray and RNA-seq data. BMC Bioinformatics 14:7. doi: 10.1186/1471-2105-14-7

11. Kang Z, Li S, Wang L, Chen Q, Hu W, Lei T, Mao Y, Zhang J, Xiang X, Wang Q, He Z, Sun T, Wang Y, Huang M, Zhang R, Chen F, Li W (2025) SYHA1813, A VEGFR and CSF1R Inhibitor, in Patients With Recurrent High-Grade Gliomas: A Multicenter, Open-Label Phase I Study. Ann Clin Transl Neurol 12:2349–2357. doi: 10.1002/acn3.70166

12. Kovats S (2015) Estrogen receptors regulate innate immune cells and signaling pathways. Cell Immunol 294:63–69. doi: 10.1016/j.cellimm.2015.01.018

13. Lan Y, Li S, Wang J, Yang X, Wang C, Huang M, Zhang R, Chen F, Li W (2025) A novel compound, SYHA1813, inhibits malignant meningioma growth directly by boosting p53 pathway activation and impairing DNA repair. Front Oncol 15:1522249. doi: 10.3389/fonc.2025.1522249

14. Liberzon A, Birger C, Thorvaldsdóttir H, Ghandi M, Mesirov JP, Tamayo P (2015) The Molecular Signatures Database (MSigDB) hallmark gene set collection. Cell Syst 1:417–425. doi: 10.1016/j.cels.2015.12.004

15. Lotsch C, Warta R, Liu F, Jungwirth G, Rommel C, Barthel M, Lamszus K, Kessler AF, Grabe N, Loehr M, Ketter R, Senft C, Maas SLN, Sievers P, Westphal M, Krieg SM, Unterberg A, Simon M, von Deimling A, Sahm F, Raleigh DR, Herold-Mende C (2025) Tumor-associated macrophages in meningiomas: a novel biomarker for poor survival outperforming the benefits of T cells. Acta Neuropathol 150:41. doi: 10.1007/s00401-025-02948-6

16. Maas SLN, Tang Y, Stutheit-Zhao E, Rahmanzade R, Blume C, Hielscher T, Zettl F, Benfatto S, Calafato D, Sill M, Benotmane JK, Yabo YA, Behling F, Suwala A, Kardo H, Ritter M, Peyre M, Sankowski R, Okonechnikov K, Sievers P, Patel A, Reuss D, Friedrich MJ, Stichel D, Schrimpf D, Van den Bosch TPP, Beck K, Wirsching H-G, Jungwirth G, Hanemann CO, Lamszus K, Etminan N, Unterberg A, Mawrin C, Remke M, Ayrault O, Lichter P, Reifenberger G, Platten M, Kacprowski T, List M, Pauling JK, Baumbach J, Milde T, Grossmann R, Ram Z, Ratliff M, Mallm J-P, Neidert MC, Bos EM, Prinz M, Weller M, Acker T, Hartmann FJ, Preusser M, Tabatabai G, Herold-Mende C, Krieg SM, Jones DTW, Pfister SM, Wick W, Kalamarides M, von Deimling A, Heiland DH, Hovestadt V, Gerstung M, Schlesner M, German “Aggressive Meningiomas” Consortium (KAM), Sahm F (2026) A microenvironment-determined risk continuum refines subtyping in meningioma and reveals determinants of machine learning-based tumor classification. Nat Genet 58:341–354. doi: 10.1038/s41588-025-02475-w

17. Nassiri F, Liu J, Patil V, Mamatjan Y, Wang JZ, Hugh-White R, Macklin AM, Khan S, Singh O, Karimi S, Corona RI, Liu LY, Chen CY, Chakravarthy A, Wei Q, Mehani B, Suppiah S, Gao A, Workewych AM, Tabatabai G, Boutros PC, Bader GD, de Carvalho DD, Kislinger T, Aldape K, Zadeh G (2021) A clinically applicable integrative molecular classification of meningiomas. Nature 597:119–125. doi: 10.1038/s41586-021-03850-3

18. Naugler WE, Sakurai T, Kim S, Maeda S, Kim K, Elsharkawy AM, Karin M (2007) Gender disparity in liver cancer due to sex differences in MyD88-dependent IL-6 production. Science 317:121–124. doi: 10.1126/science.1140485

19. Newman AM, Steen CB, Liu CL, Gentles AJ, Chaudhuri AA, Scherer F, Khodadoust MS, Esfahani MS, Luca BA, Steiner D, Diehn M, Alizadeh AA (2019) Determining cell type abundance and expression from bulk tissues with digital cytometry. Nat Biotechnol 37:773–782. doi: 10.1038/s41587-019-0114-2

20. Ning C, Xie B, Zhang L, Li C, Shan W, Yang B, Luo X, Gu C, He Q, Jin H, Chen X, Zhang Z, Feng Y (2016) Infiltrating Macrophages Induce ERα Expression through an IL17A-mediated Epigenetic Mechanism to Sensitize Endometrial Cancer Cells to Estrogen. Cancer Res 76:1354–1366. doi: 10.1158/0008-5472.CAN-15-1260

21. Ostrom QT, Price M, Neff C, Cioffi G, Waite KA, Kruchko C, Barnholtz-Sloan JS (2022) CBTRUS Statistical Report: Primary Brain and Other Central Nervous System Tumors Diagnosed in the United States in 2015-2019. Neuro Oncol 24:v1–v95. doi: 10.1093/neuonc/noac202

22. Piccolo D, Vindigni M (2026) Single-Cell-Validated Transcriptomic Proxies for the Maas Meningioma Microenvironment Risk Continuum: An NF2-Dependent Signal Attenuated Below Detectability in Bulk RNA-seq. medRxiv 2026.04.27.26351840. doi: 10.64898/2026.04.27.26351840

23. Pravdenkova S, Al-Mefty O, Sawyer J, Husain M (2006) Progesterone and estrogen receptors: opposing prognostic indicators in meningiomas. J Neurosurg 105:163–173. doi: 10.3171/jns.2006.105.2.163

24. Proctor DT, Huang J, Lama S, Albakr A, Van Marle G, Sutherland GR (2019) Tumor-associated macrophage infiltration in meningioma. Neurooncol Adv 1:vdz018. doi: 10.1093/noajnl/vdz018

25. Racle J, de Jonge K, Baumgaertner P, Speiser DE, Gfeller D (2017) Simultaneous enumeration of cancer and immune cell types from bulk tumor gene expression data. Elife 6:e26476. doi: 10.7554/eLife.26476

26. Ritchie ME, Phipson B, Wu D, Hu Y, Law CW, Shi W, Smyth GK (2015) limma powers differential expression analyses for RNA-sequencing and microarray studies. Nucleic Acids Res 43:e47. doi: 10.1093/nar/gkv007

27. Schoenfeld D (1982) Partial residuals for the proportional hazards regression model. Biometrika 69:239–241. doi: 10.1093/biomet/69.1.239

28. Teranishi Y, Miyawaki S, Nakatochi M, Okano A, Ohara K, Hongo H, Ishigami D, Sakai Y, Shimada D, Takayanagi S, Ikemura M, Komura D, Katoh H, Mitsui J, Morishita S, Ushiku T, Ishikawa S, Nakatomi H, Saito N (2023) Meningiomas in patients with neurofibromatosis type 2 predominantly comprise “immunogenic subtype” tumours characterised by macrophage infiltration. Acta Neuropathol Commun 11:156. doi: 10.1186/s40478-023-01645-3

29. Thirimanne HN, Almiron-Bonnin D, Nuechterlein N, Arora S, Jensen M, Parada CA, Qiu C, Szulzewsky F, English CW, Chen WC, Sievers P, Nassiri F, Wang JZ, Klisch TJ, Aldape KD, Patel AJ, Cimino PJ, Zadeh G, Sahm F, Raleigh DR, Shendure J, Ferreira M, Holland EC (2024) Meningioma transcriptomic landscape demonstrates novel subtypes with regional associated biology and patient outcome. Cell Genom 4:100566. doi: 10.1016/j.xgen.2024.100566

30. Thirimanne HN, Almiron-Bonnin D, Nuechterlein N, Arora S, Jensen M, Parada CA, Qiu C, Szulzewsky F, English CW, Chen WC, Sievers P, Nassiri F, Wang JZ, Klisch TJ, Aldape KD, Patel AJ, Cimino PJ, Zadeh G, Sahm F, Raleigh DR, Shendure J, Ferreira M, Holland EC (2024) Meningioma transcriptomic landscape demonstrates novel subtypes with regional associated biology and patient outcome. Cell Genom 4:100566. doi: 10.1016/j.xgen.2024.100566

31. Wu D, Smyth GK (2012) Camera: a competitive gene set test accounting for inter-gene correlation. Nucleic Acids Res 40:e133. doi: 10.1093/nar/gks461

32. Zhang Q, Guo D, Xu G, Xie R, Deng Y, Fu P, Wan J (2026) Tumor-associated macrophages suppress estrogen receptor-β expression in triple-negative breast cancer through the PI3K/AKT pathway. Exp Ther Med 31:77. doi: 10.3892/etm.2026.13072

